# Covid-19 and Excess Mortality in Medicare Beneficiaries

**DOI:** 10.1101/2021.04.07.21254793

**Authors:** Scott Greenwald, Nassib G Chamoun, Paul J Manberg, Josh Gray, David Clain, Kamal Maheshwari, Daniel I. Sessler

## Abstract

We estimated excess mortality in Medicare recipients with probable and confirmed Covid-19 infections in the general community and amongst residents of long-term care (LTC) facilities. We considered 28,389,098 Medicare and dual-eligible recipients from one year before February 29, 2020 through September 30, 2020, with mortality followed through November 30^th^, 2020. Probable and confirmed Covid-19 diagnoses, presumably mostly symptomatic, were determined from ICD-10 codes. We developed a Risk Stratification Index (RSI) mortality model which was applied prospectively to establish baseline mortality risk. Excess deaths attributable to Covid-19 were estimated by comparing actual-to-expected deaths based on historical comparisons and in closely matched cohorts with and without Covid-19. 677,100 (2.4%) beneficiaries had confirmed Covid-19 and 2,917,604 (10.3%) had probable Covid-19. 472,329 confirmed cases were community living and 204,771 were in LTC. Mortality following a probable or confirmed diagnosis in the community increased from an expected incidence of about 4% to actual incidence of 7.5%. In long-term care facilities, the corresponding increase was from 20.3% to 24.6%. The absolute increase was therefore similar at 3-4% in the community and in LTC residents. The percentage increase was far greater in the community (89%) than among patients in chronic care facilities (21%) who had higher baseline risk. The LTC population without probable or confirmed Covid-19 diagnoses experienced 38,932 excess deaths (35%) compared to historical estimates. Limitations in access to Covid-19 testing and disease under-reporting in LTC patients probably were important factors, although social isolation and disruption in usual care presumably also contributed. Remarkably, there were 31,360 fewer deaths than expected in community dwellers without probable or confirmed Covid-19 diagnoses, representing a 6% reduction. Disruptions to the healthcare system and avoided medical care were thus apparently offset by other factors, representing overall benefit. The Covid-19 pandemic had marked effects on mortality, but the effects were highly context-dependent.

## Introduction

The Covid-19 pandemic has profoundly influenced US healthcare, especially among Medicare recipients who are mostly at least 65 years old. By March 1, 2021, SARS-CoV-2, the virus responsible for Covid-19, had already infected more than 29 million Americans and more than 500,000 died following infection.(1) However, many people infected with Covid-19 are never tested or have false-negative test results; consequently, the true toll of Covid-19 remains uncertain. Furthermore, while the clinical course is sometimes apparent, Covid-19 also kills people by worsening chronic conditions, with those deaths often being attributed to other causes. Especially early in the pandemic, due to limited testing availability, it was difficult to differentiate deaths caused by Covid-19 from those that may have occurred naturally due to underlying health conditions. It is thus apparent that many people who died consequent to Covid-19 infections may not have been diagnosed with the condition or may have died due to underlying causes.

Several teams have estimated “excess” mortality due to Covid-19 by comparing weekly observed death totals with those that occurred in a prior year. For example, Chen et al estimated that from March 1 through August 22, 2020, 146,557 deaths were recorded in California, with an estimated 19,806 (95% CI: 16,364, 23,210) deaths in excess of those predicted by historical trends.(2) Similarly, Faust et al estimated that from March 1, 2020, to July 31, 2020, a total of 76,088 all-cause deaths occurred among US adults aged 25 to 44 years, which was 11,899 more than the expected 64,189 deaths based on a previous year (incident rate ratio, 1.19 [95% CI, 1.14-1.23]).(3) Rossen et al estimated excess mortality from January 26 through October 3 to have *decreased* 2% for the youngest subjects (aged <25 years) but increased 14.4% for those 45-64 years, 21.1% for those 65-74 years, 21.5% for those 75-84 years, and 14.7% in subjects ≥85 years old.(4) These reports suggest that all-cause mortality in the first six months of the pandemic increased by about 15-20%.. However, historical comparisons do not account for risk at an individual level which may be useful to determine true excess mortality.

On a broad population basis, many risk factors for Covid-19 are now well recognized. For example, the CDC identifies eleven conditions that augment risk for severe forms of Covid-19.(5) Chronic conditions such as cancer and dementia are reported to be among the most important contributors.(6-8) It is clear that older members of the population are at special risk, although to some extent age may be a surrogate for accumulated comorbidities. However, it is difficult to extrapolate from population risk to individual risk since many people exhibit various combinations of risk factors for Covid-19 mortality, and individual risks attributable to each condition are not necessarily additive. A robust model that considers relevant individual conditions and predicts mortality risk from Covid-19 infections would therefore be valuable.

Numerous groups have proposed individual risk models based on clinical outcomes in various populations studied early in the pandemic, but a consensus model has yet to emerge.(9-11) From a practical perspective, prediction models based on readily available administrative data (e.g., ICD-10 codes) will be most useful since more granular information extracted from clinical health records are neither universally available nor easy to obtain. Our primary goal was therefore to estimate excess risk-specific mortality in people with probable and confirmed Covid-19 infections.

An additional consequence of the Covid-19 pandemic has been public health quarantines that have severely disrupted healthcare delivery. The virus may therefore also have caused indirect mortality because patients with chronic diseases and acute exacerbations avoided seekin care due to fear of infection or because health services were overwhelmed or otherwise limiting access. Furthermore, stress related to isolation could increase medical morbidity and provoke suicide and substance abuse. In contrast, some causes of death such as accidents and homicides may have diminished. The extent to which delayed and disrupted healthcare for non-Covid-related conditions, along with pandemic-related behavioral changes, contribute to mortality remains unclear. Our secondary goal was therefore to estimate whether changes in mortality occurred in people without probable or confirmed Covid-19 infections.

Because Covid-19 is especially lethal in older people, we considered stratification by various age ranges for both our primary and secondary analyses. We also separately considered people residing in the community from those in long term care facilities, who are expected to have a higher baseline mortality risk and thus may be especially susceptible to Covid-19 infections.

## Methods

Data analysis was conducted on the Center for Medicare and Medicaid Services (CMS) Research Identifiable File (RIF) data using SAS Enterprise Guide (Version 7.15) under a special Data Use Agreement (DUA). This project was determined to be exempt from informed consent requirements by the New England Institutional Review Board. Final data analysis of the full cohort was conducted from January 10 to March 11, 2021. This study followed the Strengthening the Reporting of Observational Studies in Epidemiology (<u>STROBE</u>) reporting guideline for cohort studies.(12)

Individual subject data used for our analysis are available to certain stakeholders as allowed by federal regulations and CMS policy. Requests for access to data to replicate these findings require an approved research protocol and DUA with CMS. For more information, contact the Research Data Assistance Center (ResDAC, http://www.resdac.org).

Mortality predictions and outcomes were referenced to an anchor date of Feb 29, 2020, just before the initial wave of documented Covid-19 cases and the week when the first case of potential community spread Covid-19 was reported by CDC. We recognize that undiagnosed cases may have occurred previously, but it is unlikely that there were many.

We used full Medicare fee-for-service and dual eligible (Medicaid and Medicare) files one year before the anchor date through September 30, 2020, with mortality outcomes reported through November 30^th^, 2020 for the primary analysis of Covid-19 outcomes. We identified beneficiaries with confirmed Covid-19 diagnoses consistent with CMS guidance using ICD-10-CM codes for Covid-19 (B97.29 before April 1, 2020 and U07.1 thereafter) as a primary or secondary diagnosis between March 1, 2020 and September 30, 2020.(13) Probable Covid-19 infection cases were identified using ICD-10-CM codes consistent with the CDC guidance (Z20.828) and WHO recommendations (U07.2).(14, 15) Presumably most subjects with Covid-19 diagnoses were symptomatic, although some may have been tested because of risk or exposure.

Demographic characteristics (age, sex, ethnicity, location of care, zip code derived measures) and Medicare coverage information (dates of coverage, enrollment history) were extracted for each subject. Beneficiaries were classified as belonging to the long-term care/skilled nursing facility (LTC/SNF) cohort if their claims history indicated that they had received services in a LTC/SNF setting at any time in February 2020. The remaining beneficiaries were designated as community dwelling.

We included all Medicaid and Medicare participants alive as of the study anchor date (N=65,310,173). We excluded beneficiaries who had: 1) ages outside 18-99 years (214,767); 2) non-continuous coverage of Medicare Part A or B (12,445,567) or any Medicare Part C coverage (23,633,391) in the year before the Feb 29, 2020 anchor date; 3) missing data for any variable used in the analysis (509,932); and, inconsistent data (117,418). The resulting 28,389,098 beneficiaries included 677,100 with confirmed and 2,917,604 with probable diagnosis of Covid-19 (**S1 Fig**). Mortality was assumed to have occurred on the date-of-death listed in the CMS Common Medicare Environment which is continuously updated from various sources including the Social Security Administration.

### Risk Stratification

We used an adaptation of the Risk Stratification Index (RSI) to predict nine-month mortality using the prior year’s administrative claims. A model was developed using the full fee-for-service 2018 population for training, with prospective validation of performance on the 2019 dataset as previously described.(16) More detailed descriptions of the model and prospective testing performance results are provided in **S1 File** and **S2 Fig**, respectively. The resulting RSI model was then prospectively applied to all eligible Medicare or dual-eligible beneficiaries as of February 29, 2020 to derive individual RSI scores as of that date — that is, before Covid-19 infections were confirmed in the United States.

For comparative purposes, a second model was similarly developed to predict nine-month mortality from the presence of 27 individual chronic conditions as defined by CMS.(17) Specifically, logistic regression (stepwise selection using p-in of 10^−3^, p-out of 10^−2^) was used to select significant predictors from a pool of candidate features (i.e., 27 chronic conditions, age, sex and dual-enrollment status) to create a predictive model using the same training and prospective testing populations described in the previous paragraph. Performance using baseline RSI values or the individual chronic conditions model as predictors of outcomes in 2020 were then tested prospectively (**S3 Fig)**.

### Analysis and Selection of Study Cohorts

We conducted a progression of complementary inquiries. To test our primary hypothesis, we first identified the main study cohorts of beneficiaries with diagnoses of probable or confirmed Covid-19, and then subdivided them based on location of service (community or LTC/SNF) as of February 29, 2020. Within each cohort, we determined 9-month mortality between the anchor date and November 30, 2020.

Our goal was to first define associations between baseline demographic characteristics and health status as characterized by RSI with the risk of mortality following a Covid-19 diagnosis in the overall at-risk population and in pre-defined subpopulations. We initially compared differences in mean baseline RSI scores between survivors and non-survivors, then used univariable and multivariable regression modeling to estimate the relative importance of baseline demographic factors, chronic conditions, and RSI scores as independent predictors of mortality. A similar analysis was conducted to identify risk factors associated with a confirmed diagnosis of Covid-19. We also determined the association between RSI and observed mortality by beneficiary age group and location.

### Estimation of Expected Mortality

Two independent methods were used to estimate expected 2020 mortality in our study population. A historical comparison allowed us to compare year-over-year changes in mortality in Medicare recipients and thus characterize overall effects of Covid-19 and quarantine-induced restrictions in healthcare access on mortality. A case-matched analysis provided an alternate estimate of Covid-19-related excess mortality within the 9 months of 2020 that we considered.

### Historical Comparison

As in previous studies,(3, 4) the first approach estimated expected 2020 mortality figures from historical records. The daily observed mortality for the Medicare population from 2017-2019 was used to prepare a model with optimal fit to capture seasonality and account for annual trends using a three-year moving average adjustment (**S2 Fig**). This approach better estimates expected mortality than relying on a single year-over-year comparison because the model better captures year-to-year fluctuations consequent to severity of yearly influenza outbreaks and other factors. We calculated predicted mortality for each individual and designated the sum as the historically expected mortality in each subpopulation. Excess deaths thus equaled the difference between observed 2020 deaths and the historical projection of expected deaths (actual minus expected).

### Case Matching, Digital Twins

A second method used case matching or “digital twinning” to estimate excess mortality in exposed subjects compared to concurrent controls who had closely matched health profiles. Beneficiaries receiving a diagnosis of probable or confirmed Covid-19 were pairwise exactly matched 1:1 on Feb 29, 2020 with beneficiaries without a Covid-19 diagnosis based on sex, age (within 1-year), ethnicity, location of services in Feb 2020 (community or LTC/SNF), along with RSI as a propensity matching factor (within 0.1%). Because the eligible Medicare population is large, we successfully matched almost the entire infected population. Excess deaths were estimated as the difference between the observed number of deaths in probable or confirmed Covid-19 subjects and their matched non-Covid-19 digital twins over the concurrent period. Matching may be more reliable than the historical comparison for estimating true excess mortality because it better accounts for population variation over time and accounts for the impact of substantial disruptions in public health and everyday life caused by the pandemic restrictions in 2020.

### Statistical Analysis

We used SAS Enterprise Guide (Version 7.15) to build models and conduct analyzes. We excluded patients who lacked any variable used in our analyses including birth date, sex, race, Medicaid enrollment status, zip code, and baseline RSI. Additionally, we excluded subjects whose records had inconsistent values among source files containing similar variables such as birth date and sex.

Patient demographic and clinical characteristics are summarized descriptively. Baseline characteristics were compared using *t* or χ^2^ tests, as appropriate. Mortality rates within the study period post-Covid-19 diagnosis are reported as odds ratios with 95% confidence intervals. P values <0.05 defined statistical significance for both the primary and secondary outcomes. No adjustments were made for multiple comparisons. Sample size requirements were not estimated *a priori* because the intention was to include all qualifying 2020 beneficiaries available in the 100% nationwide Medicare files.

## Results

### Patient characteristics and outcomes

As of Feb 29, 2020, a total of 28,389,098 Medicare or dual eligible beneficiaries met inclusion criteria for this study. Among them, 677,100 (2.4%) beneficiaries had a diagnosis of confirmed Covid-19 and 2,917,604 (10.3%) had a diagnosis of probable Covid-19 during the study period (**S1 Fig**). Among the confirmed cases, 472,329 were in the Community group while 204,771 received care in a long-term care setting.

**Table1** and **Table 2** compare demographic and clinical profiles for various subgroups. Compared to survivors, patients who died after a Covid-19 diagnosis were older, more often male, not white, received Medicaid, lived in zip-codes associated with lower median income, received services in February 2020 in a long-term care facility, and had higher baseline risk of mortality as defined by RSI. Age and baseline RSI scores were both strongly related to risk of infection and adverse Covid-19 outcomes. Residence in a LTC/SNF location and presence of end-stage renal disease were strong risk factors for acquiring a confirmed diagnosis of Covid-19 (**S4 Fig**). As shown in **S5 Fig**, RSI scores were associated with increasing mortality in a consistent rank ordered manner across each age group, thereby suggesting that RSI provides a significant and sensitive measure of co-morbidities and mortality risk that is independent of age.

**Table 1.** Comparison of baseline demographic characteristics of all study populations.

| Characteristic | Group Size (N,%) |  |  |  |  |  |  |  |  |
| --- | --- | --- | --- | --- | --- | --- | --- | --- | --- |
|  | All Beneficiaries<br>(28,389,098) | COVID Confirmed<br>(677,100) | Pr < Z | Died<br>(131,460) | Survived<br>(545,640) | COVID Probable<br>(2,917,604) | Pr < Z | Died<br>(214,602) | Survived<br>(2,703,002) |
| **Age Group** |  |  |  |  |  |  |  |  |  |
| 18-55 | 1,938,606<br>(6.8) | 43,637<br>(6.4) | <.0001 | 3,070<br>(2.3) | 40,567<br>(7.4) | 210,430<br>(7.2) | <.0001 | 6,155<br>(2.9) | 204,275<br>(7.6) |
| 56-65 | 1,912,571<br>(6.7) | 57,655<br>(8.5) | <.0001 | 7,630<br>(5.8) | 50,025<br>(9.2) | 224,790<br>(7.7) | <.0001 | 12,503<br>(5.8) | 212,287<br>(7.9) |
| 66-70 | 6,900,760<br>(24.3) | 118,536<br>(17.5) | <.0001 | 12,145<br>(9.2) | 106,391<br>(19.5) | 643,015<br>(22.0) | <.0001 | 22,527<br>(10.5) | 620,488<br>(23.0) |
| 71-75 | 6,707,830<br>(23.6) | 123,535<br>(18.2) | <.0001 | 16,740<br>(12.7) | 106,795<br>(19.6) | 645,540<br>(22.1) | <.0001 | 30,035<br>(14.0) | 615,505<br>(22.8) |
| 76-80 | 4,688,185<br>(16.5) | 103,925<br>(15.3) | <.0001 | 20,067<br>(15.3) | 83,858<br>(15.4) | 476,297<br>(16.3) | <.0001 | 34,007<br>(15.8) | 442,290<br>(16.4) |
| 81-85 | 3,140,391<br>(11.1) | 88,874<br>(13.1) | <.0001 | 22,686<br>(17.3) | 66,188<br>(12.1) | 333,813<br>(11.4) | <.0001 | 36,602<br>(17.1) | 297,211<br>(11.0) |
| 86-90 | 1,914,030<br>(6.7) | 74,295<br>(11.0) | <.0001 | 23,529<br>(17.9) | 50,766<br>(9.3) | 224,030<br>(7.7) | <.0001 | 35,949<br>(16.8) | 188,081<br>(7.0) |
| 91-95 | 947,362<br>(3.3) | 50,455<br>(7.5) | <.0001 | 18,665<br>(14.2) | 31,790<br>(5.8) | 125,819<br>(4.3) | <.0001 | 27,433<br>(12.8) | 98,386<br>(3.6) |
| 96-99 | 239,363<br>(0.8) | 16,188<br>(2.4) | <.0001 | 6,928<br>(5.3) | 9,260<br>(1.7) | 33,870<br>(1.2) | <.0001 | 9,391<br>(4.4) | 24,479<br>(0.9) |
| **Sex** |  |  |  |  |  |  |  |  |  |
| Female | 15,538,151<br>(54.7) | 385,062<br>(56.9) | <.0001 | 68,726<br>(52.3) | 316,336<br>(58.0) | 1,655,208<br>(56.7) | <.0001 | 110,919<br>(51.7) | 1,544,289<br>(57.1) |
| Male | 12,850,947<br>(45.3) | 292,038<br>(43.1) | <.0001 | 62,734<br>(47.7) | 229,304<br>(42.0) | 1,262,396<br>(43.3) | <.0001 | 103,683<br>(48.3) | 1,158,713<br>(42.9) |
| **Race/Ethnicity** |  |  |  |  |  |  |  |  |  |
| American Indian/Alaskan Native | 161,358<br>(0.6) | 5,194<br>(0.8) | <.0001 | 1,171<br>(0.9) | 4,023<br>(0.7) | 17,312<br>(0.6) | <.0001 | 1,380<br>(0.6) | 15,932<br>(0.6) |
| Asian/Pacific Islander | 766,965<br>(2.7) | 16,894<br>(2.5) | <.0001 | 3,679<br>(2.8) | 13,215<br>(2.4) | 57,742<br>(2.0) | <.0001 | 4,270<br>(2.0) | 53,472<br>(2.0) |
| Black | 2,308,293<br>(8.1) | 97,484<br>(14.4) | <.0001 | 21,394<br>(16.3) | 76,090<br>(13.9) | 263,726<br>(9.0) | <.0001 | 20,962<br>(9.8) | 242,764<br>(9.0) |
| Hispanic | 1,577,063<br>(5.6) | 65,953<br>(9.7) | <.0001 | 12,760<br>(9.7) | 53,193<br>(9.7) | 151,585<br>(5.2) | <.0001 | 9,898<br>(4.6) | 141,687<br>(5.2) |
| Non-Hispanic White | 22,806,998<br>(80.3) | 477,687<br>(70.5) | <.0001 | 90,660<br>(69.0) | 387,027<br>(70.9) | 2,352,811<br>(80.6) | <.0001 | 175,365<br>(81.7) | 2,177,446<br>(80.6) |
| Other | 228,402<br>(0.8) | 5,164<br>(0.8) | 0.0001 | 1,004<br>(0.8) | 4,160<br>(0.8) | 20,356<br>(0.7) | <.0001 | 1,272<br>(0.6) | 19,084<br>(0.7) |
| Unknown | 540,019<br>(1.9) | 8,724<br>(1.3) | <.0001 | 792<br>(0.6) | 7,932<br>(1.5) | 54,072<br>(1.9) | <.0001 | 1,455<br>(0.7) | 52,617<br>(1.9) |
| **Low Income/Disabled Status** |  |  |  |  |  |  |  |  |  |
| Low Income or Disabled | 8,222,944<br>(29.0) | 329,258<br>(48.6) | <.0001 | 69,107<br>(52.6) | 260,151<br>(47.7) | 982,097<br>(33.7) | <.0001 | 82,852<br>(38.6) | 899,245<br>(33.3) |
| Not Low Income/Disabled | 20,166,154<br>(71.0) | 347,842<br>(51.4) | <.0001 | 62,353<br>(47.4) | 285,489<br>(52.3) | 1,935,507<br>(66.3) | <.0001 | 131,750<br>(61.4) | 1,803,757<br>(66.7) |
| **Median Household Income (Imputed from Zip code) (Quintile)** |  |  |  |  |  |  |  |  |  |
| 20-40% | 5,674,738<br>(20.0) | 121,354<br>(17.9) | <.0001 | 23,044<br>(17.5) | 98,310<br>(18.0) | 520,937<br>(17.9) | <.0001 | 42,043<br>(19.6) | 478,894<br>(17.7) |
| 40-60% | 5,679,860<br>(20.0) | 122,059<br>(18.0) | <.0001 | 23,585<br>(17.9) | 98,474<br>(18.0) | 533,097<br>(18.3) | <.0001 | 41,889<br>(19.5) | 491,208<br>(18.2) |
| 60-80% | 5,675,343<br>(20.0) | 122,827<br>(18.1) | <.0001 | 23,660<br>(18.0) | 99,167<br>(18.2) | 599,106<br>(20.5) | <.0001 | 42,578<br>(19.8) | 556,528<br>(20.6) |
| Highest 20% | 5,679,506<br>(20.0) | 152,382<br>(22.5) | <.0001 | 28,850<br>(21.9) | 123,532<br>(22.6) | 725,509<br>(24.9) | <.0001 | 42,077<br>(19.6) | 683,432<br>(25.3) |
| Lowest 20% | 5,679,651<br>(20.0) | 158,478<br>(23.4) | <.0001 | 32,321<br>(24.6) | 126,157<br>(23.1) | 538,955<br>(18.5) | <.0001 | 46,015<br>(21.4) | 492,940<br>(18.2) |

| **Community vs LTC/SNF** |  |  |  |  |  |  |  |  |  |
| --- | --- | --- | --- | --- | --- | --- | --- | --- | --- |
| Community | 27,400,582<br>(96.5) | 472,329<br>(69.8) | <.0001 | 69,488<br>(52.9) | 402,841<br>(73.8) | 2,676,502<br>(91.7) | <.0001 | 166,800<br>(77.7) | 2,509,702<br>(92.8) |
| In Long term<br>care Facility or<br>SNF | 988,516<br>(3.5) | 204,771<br>(30.2) | <.0001 | 61,972<br>(47.1) | 142,799<br>(26.2) | 241,102<br>(8.3) | <.0001 | 47,802<br>(22.3) | 193,300<br>(7.2) |
| **RSI (9mo)** |  |  |  |  |  |  |  |  |  |
| MEAN<br>(SD) | 0.032<br>(.071) | 0.096<br>(.127) | <.0001 | 0.181<br>(.153) | 0.075<br>(.110) | 0.052<br>(.095) | <.0001 | 0.168<br>(.155) | 0.042<br>(.082) |
| **Mortality**<br>**(Through Nov**<br>**30)** |  |  |  |  |  |  |  |  |  |
| Died | 1,045,326<br>(3.7) | 131,460<br>(19.4) | <.0001 | 131,460<br>(100.0) | 0<br>(0.0) | 214,602<br>(7.4) | <.0001 | 214,602<br>(100.0) | 0<br>(0.0) |
| Survived | 27,343,772<br>(96.3) | 545,640<br>(80.6) | <.0001 | 0<br>(0.0) | 545,640<br>(100.0) | 2,703,002<br>(92.6) | <.0001 | 0<br>(0.0) | 2,703,002<br>(100.0) |

\*p-values represent the level of significance comparing attributes between Confirmed and Probable Covid infected populations relative to all Beneficiaries.
Confirmed Covid-19 cases were identified consistent with CMS guidance using ICD-10-CM codes for Covid-19 (B97.29 before April 1, 2020 and U07.1 thereafter) as a primary or secondary diagnosis between March 1, 2020 and September 30, 2020.<sup>12</sup> Probable Covid-19 infection cases were identified using ICD-10-CM codes consistent with the CDC guidance (Z20.828) and WHO recommendations (U07.2).<sup>13,14</sup> The baseline risk of 9-month mortality defined by the Risk Stratification Index (RSI) calculated on February 29, 2020 was 3.2% in the entire population and significantly higher among those who died compared to those who survived.

**Table 2.**
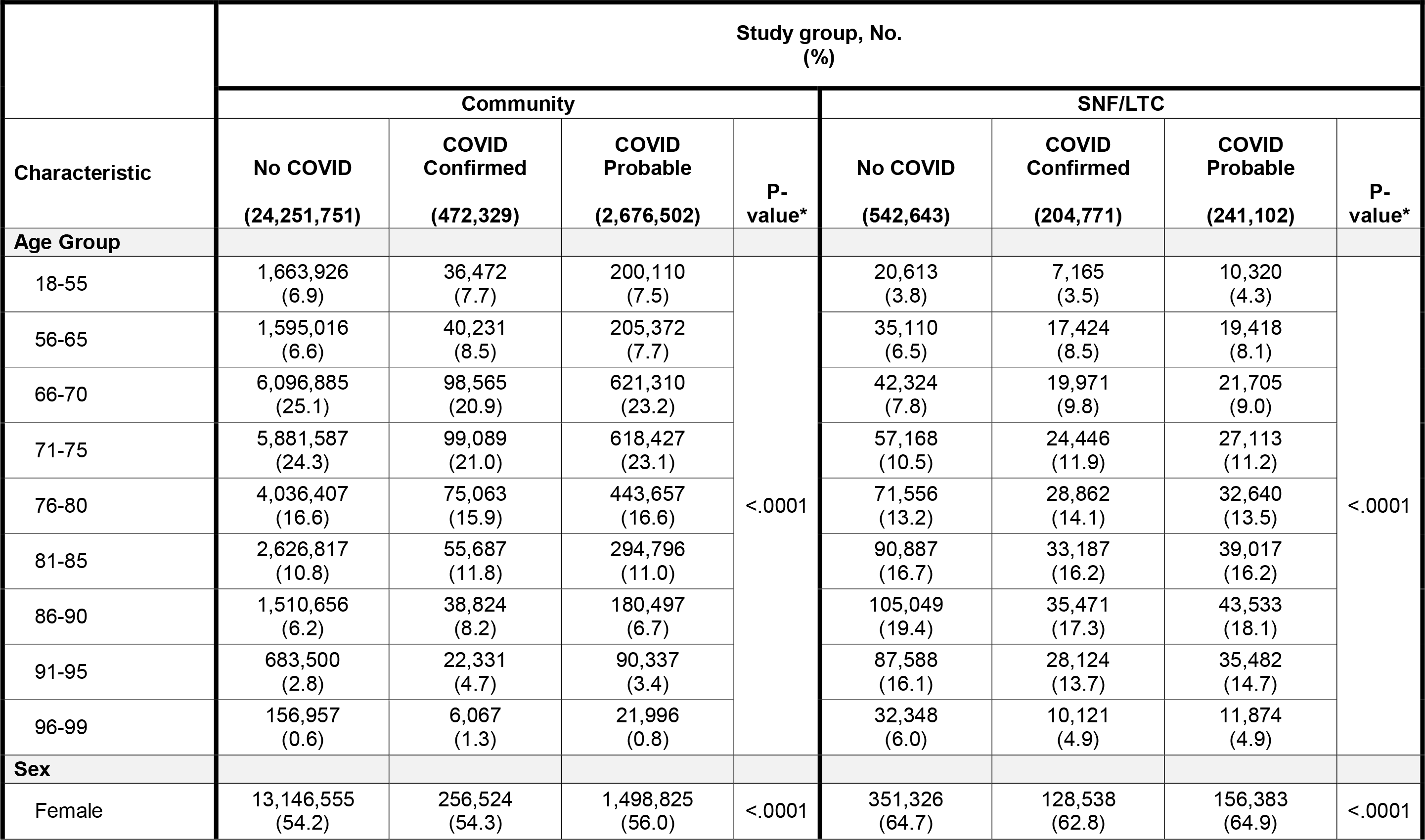
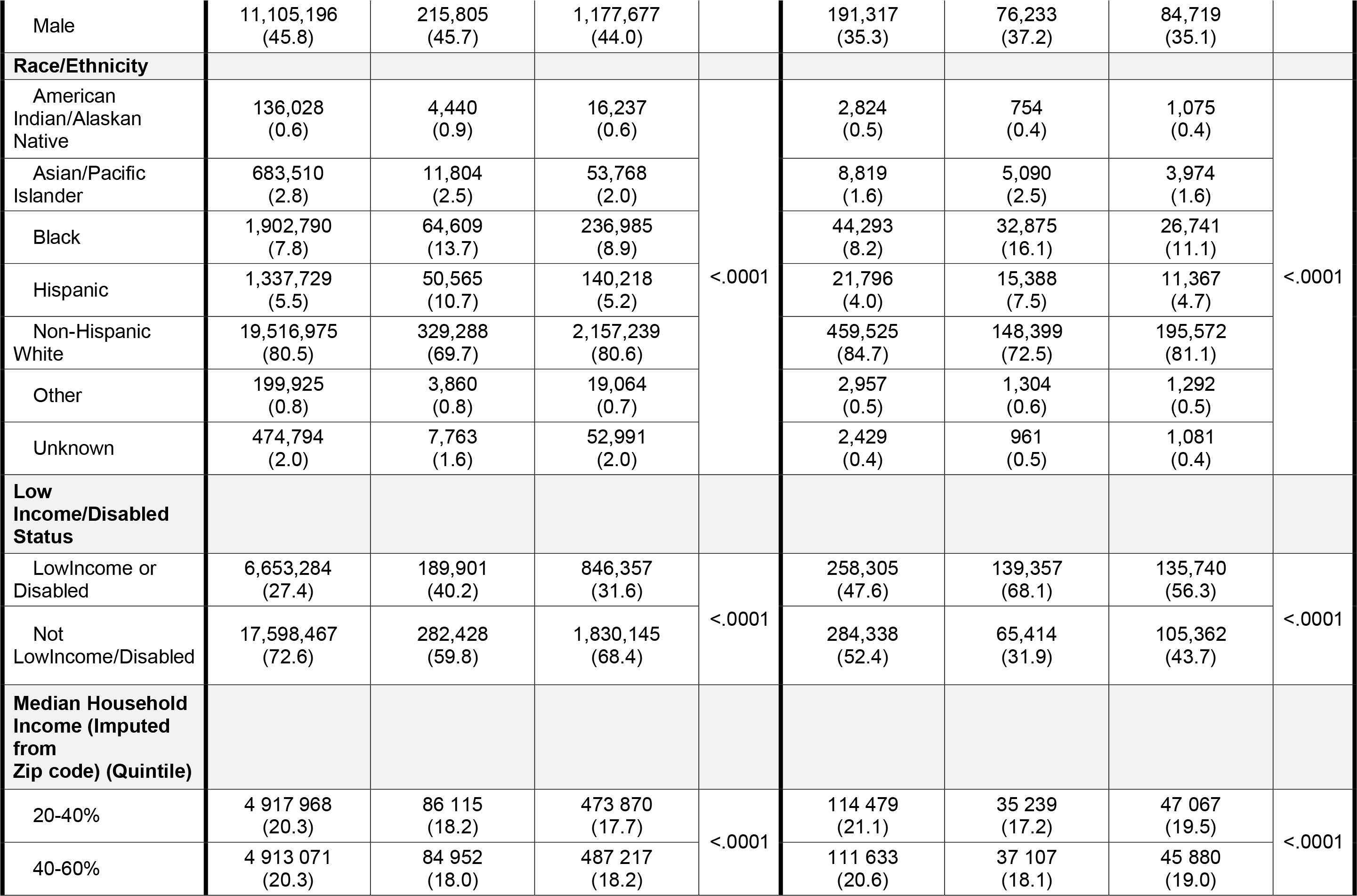
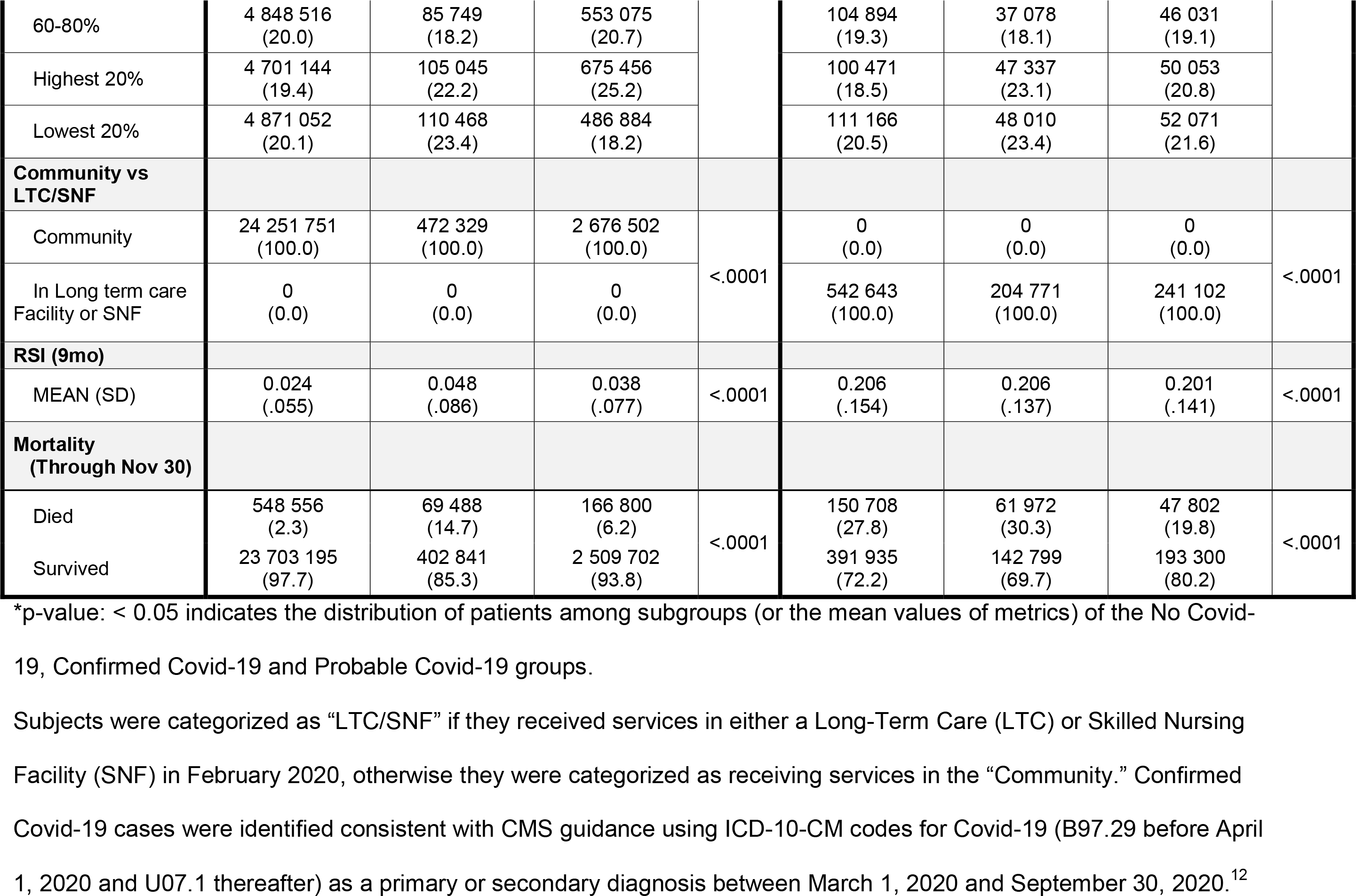
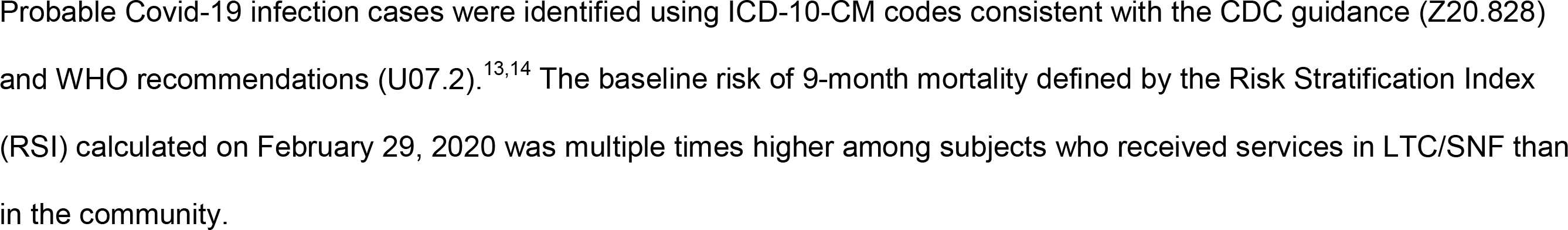
Comparison of baseline demographic characteristics for No Covid-19, Confirmed Covid-19 and Probable Covid-19 populations in the Community and LTC/SNF subgroups.

### Mortality Risk Prediction

**Fig 1** presents the relative importance of factors that contributed to mortality in both univariable and multivariable models. Quintiles of RSI, age, LTC/SNF services status, sex, and race were the factors most associated with relative risk of mortality. Status of lung cancer and end-stage renal disease appear to carry meaningful incremental risk after adjustment. Mortality prediction models based primarily on baseline RSI levels performed better than models based on the presence of individual chronic conditions for predicting mortality risk (**S3 Fig**). Case matching identified a cohort of beneficiaries from the general population who were closely matched with subjects who had a diagnosis of probable or confirmed Covid-19 based on their RSI scores as of Feb 29, 2020 (**Table 3**).

**Table 3.**
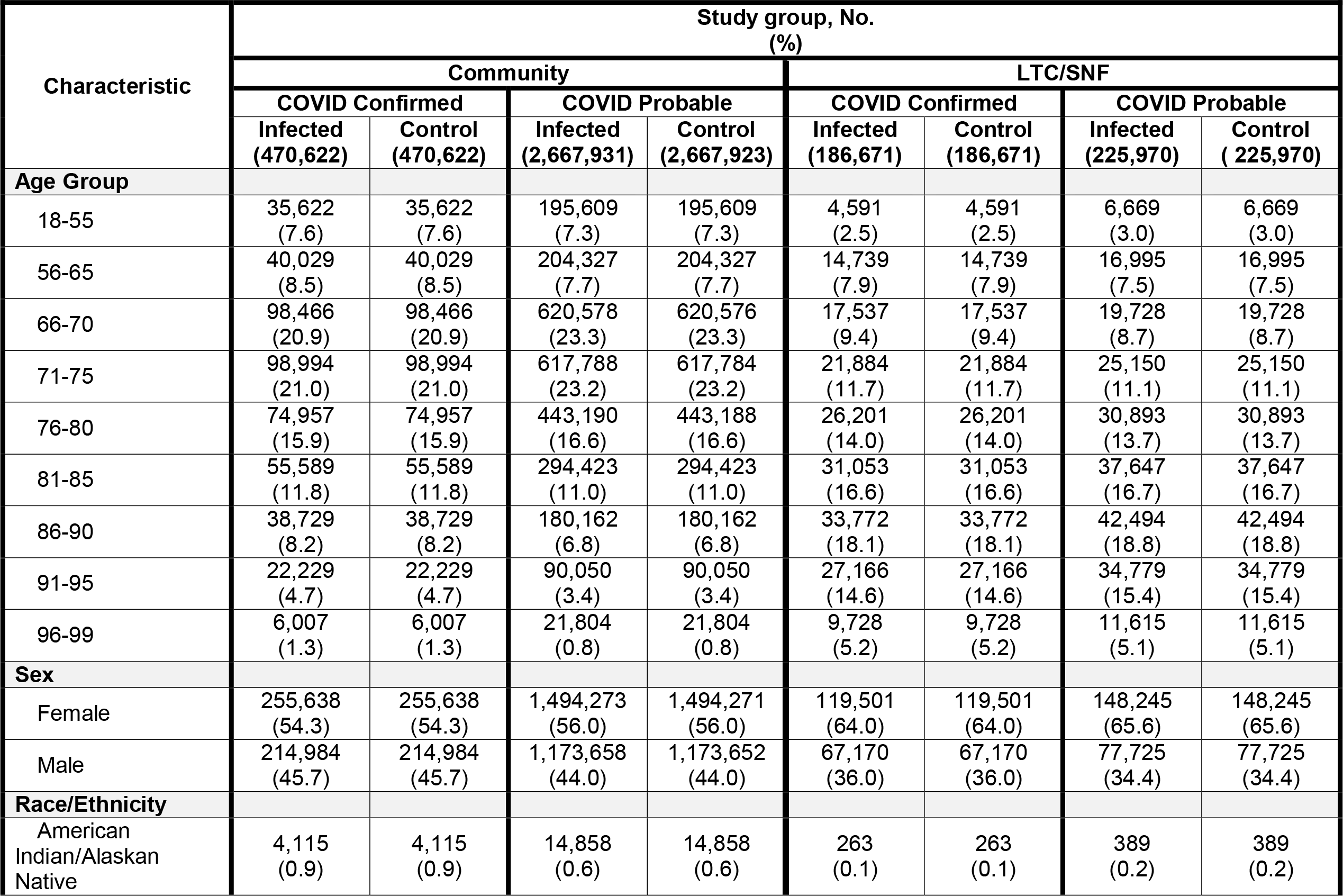
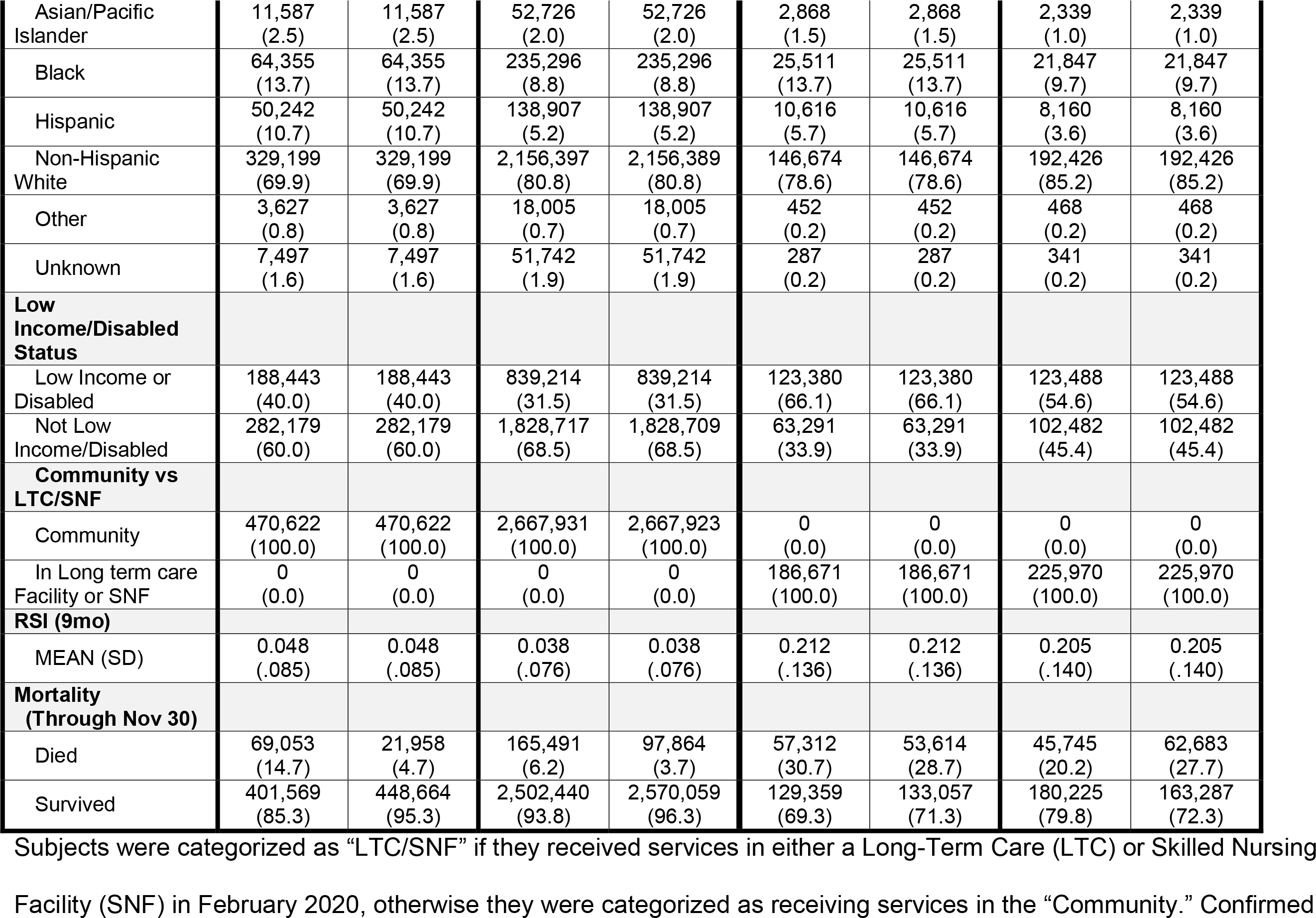
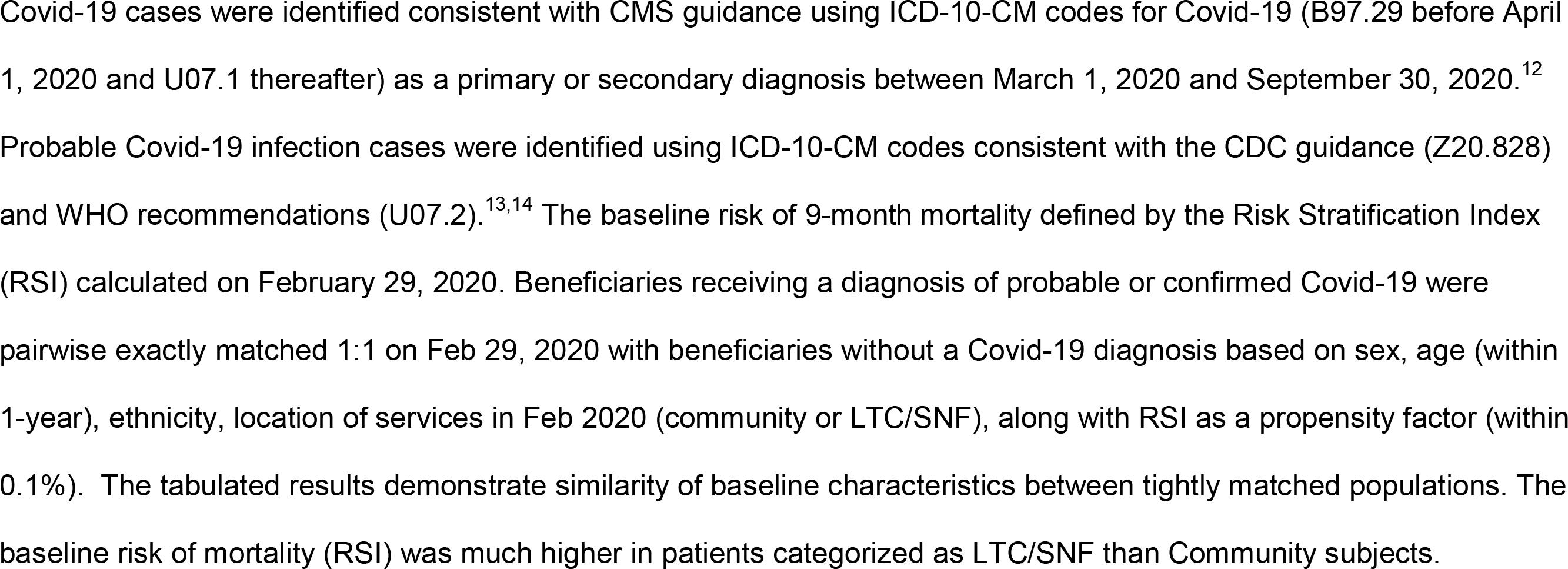
Comparison of matching characteristics for Covid-19 subjects versus non-Covid-19 controls in the Community and LTC/SNF subgroups.

**Fig 1.**
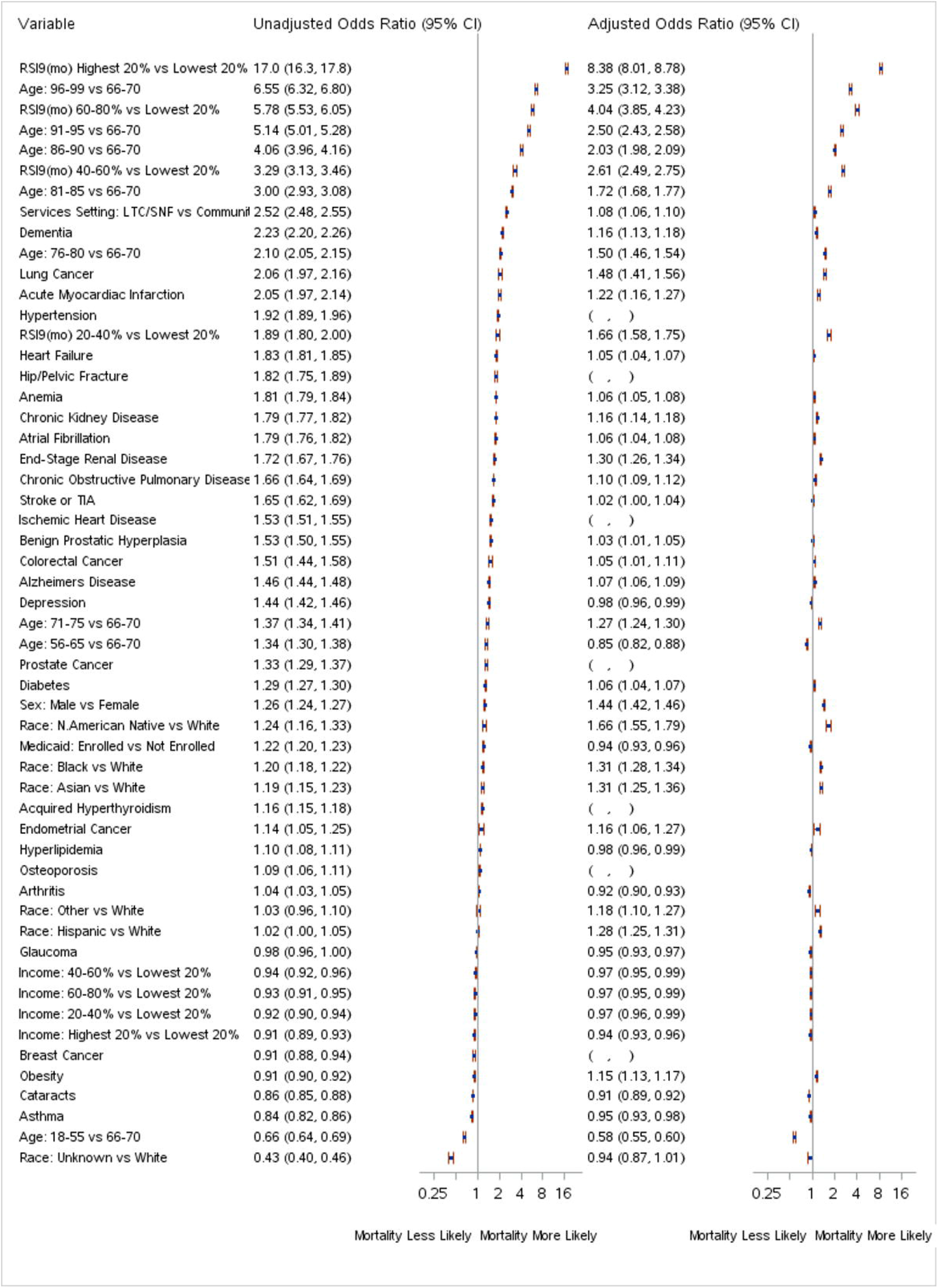
Forest plot showing the relative risk and 95% CI of significant predictors of mortality of subjects with confirmed Covid-19. Confirmed Covid-19 cases were identified consistent with CMS guidance using ICD-10-CM codes for Covid-19 (B97.29 before April 1, 2020 and U07.1 thereafter) as a primary or secondary diagnosis between March 1, 2020 and September 30, 2020.^12^ Subjects were categorized as “LTC/SNF” if they received services in either a Long Term Care (LTC) or Skilled Nursing Facility (SNF) in February 2020, otherwise they were categorized as receiving services in the “Community.” Predictors were assessed at baseline (February 29, 2020) and include quintiles of Risk Stratification Index (RSI), presence of chronic conditions, location of services (LTC/SNF vs Community), and demographic variables (i.e., age, sex, race, and quintiles of median household income imputed by zip code according to 2015 Census data.) Variables not remaining in the adjusted model are indicated by the presence of empty parenthesis under the adjusted odds ratio. RSI, age, and location of services were the strongest (unadjusted) predictors of mortality. RSI and age remain strong predictors following adjustment; however, risks associated with having chronic conditions were typically reduced when adjusted by the presence of RSI and other factors. Status of Lung cancer and end-stage renal disease appear to carry meaningful incremental risk after adjustment.

### Excess Mortality Estimates

The distribution of observed and expected mortality by diagnosis, category, and location of care is presented in **Fig 2**. As expected, subjects with high baseline mortality risk in the LTC/SNF cohort had actual mortality that exceeded all other groups. Those with confirmed Covid-19 showed similarly increased mortality above expected levels in both the LTC/SNF and community setting. Among community dwelling subjects, mortality also exceeded expected risk in subjects with possible Covid-19.

**Fig 2.**
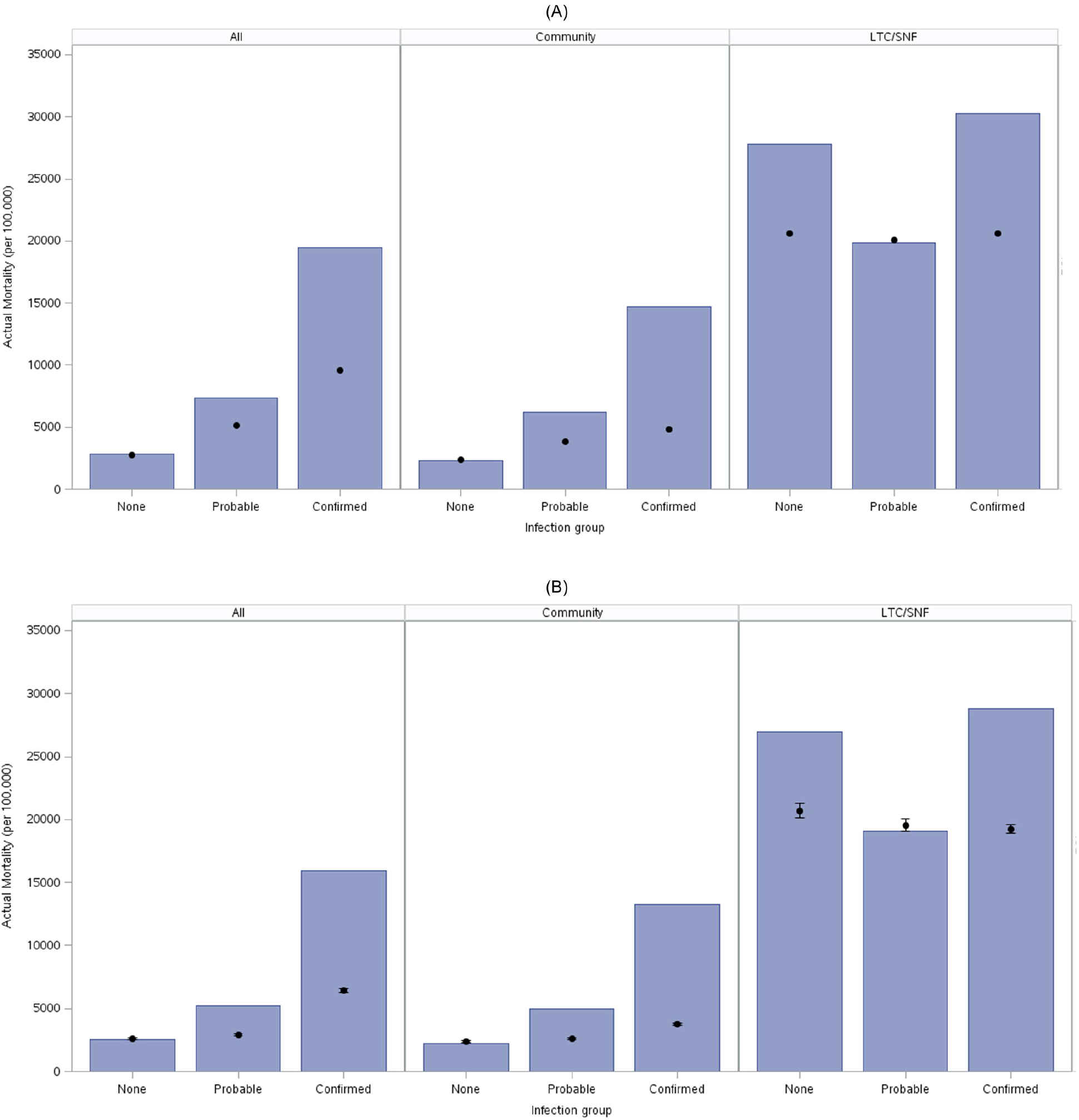
Actual/Expected mortality plot for No Covid-19, Probable Covid-19, and Confirmed Covid −19 cohorts in community, LTC/SNF, and combined analysis. Panels A and B display actual and expected mortality (per 100,000 people) calculated using different methods for Medicare subjects grouped by infection status and location of services. Confirmed Covid-19 cases were identified consistent with CMS guidance using ICD-10-CM codes for Covid-19 (B97.29 before April 1, 2020 and U07.1 thereafter) as a primary or secondary diagnosis between March 1, 2020 and September 30, 2020.^12^ Probable Covid-19 infection cases were identified using ICD-10-CM codes consistent with the CDC guidance (Z20.828) and WHO recommendations (U07.2).^13,14^ Subjects were categorized as “LTC/SNF” if they received services in either a Long Term Care (LTC) or Skilled Nursing Facility (SNF) in February 2020, otherwise they were categorized as receiving services in the “Community.” Estimated mortality using RSI (A) provides estimates consistent with actual mortality of historical controls (B).

There was an estimated excess of 130,702 (historical comparison method) or 101,482 (case matching method) deaths attributable to probable or confirmed Covid-19 across the full population in the 9 months of 2020 that we considered. In the matched analysis, half the deaths (50,793) occurred in patients with a confirmed diagnosis of Covid-19 and half (50,689) occurred in those with a probable Covid-19 diagnosis. In contrast, 31,360 fewer subjects without a Covid-19 diagnosis died than expected, representing a 6% mortality reduction (**Table 4**).

**Table 4.**
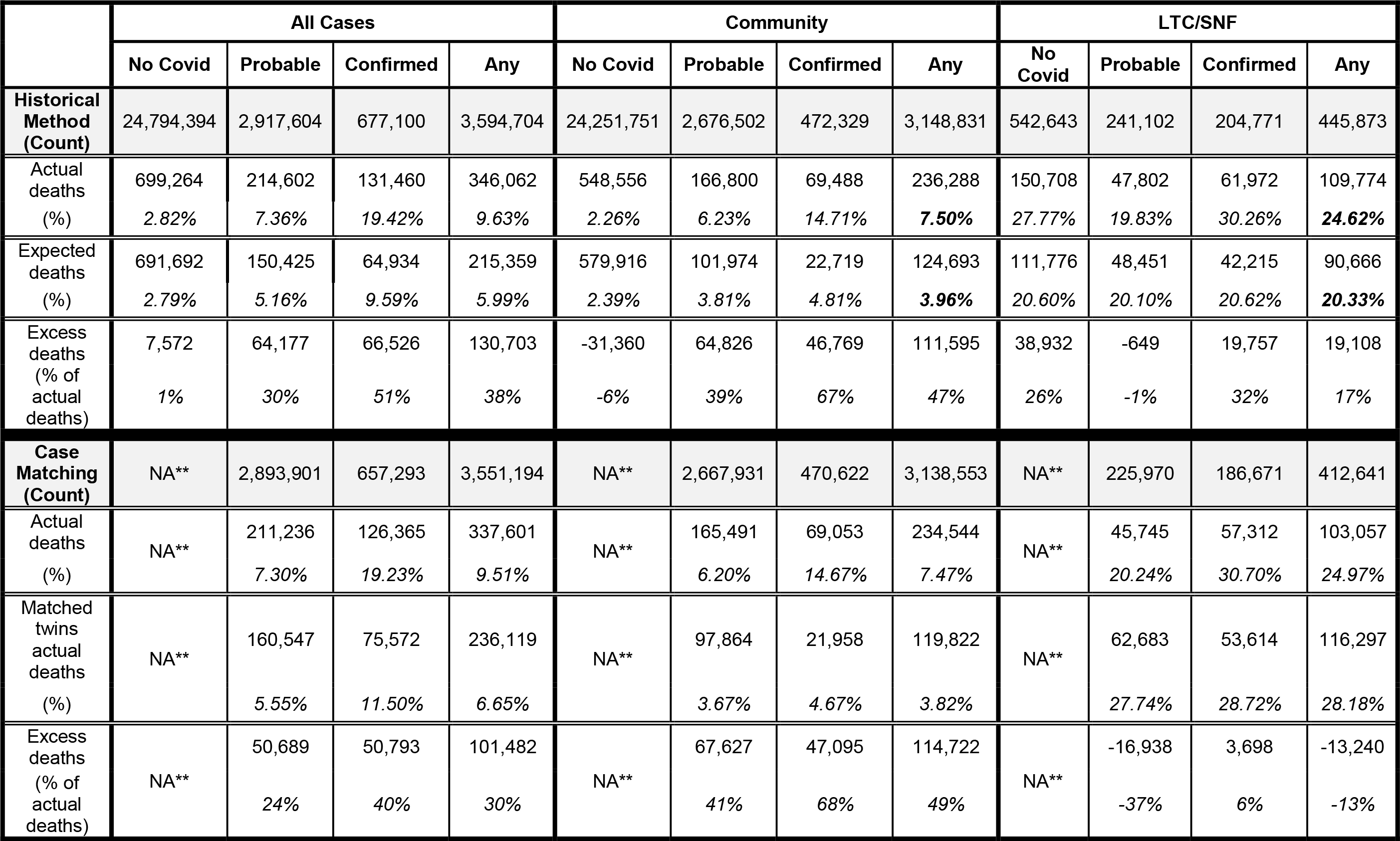
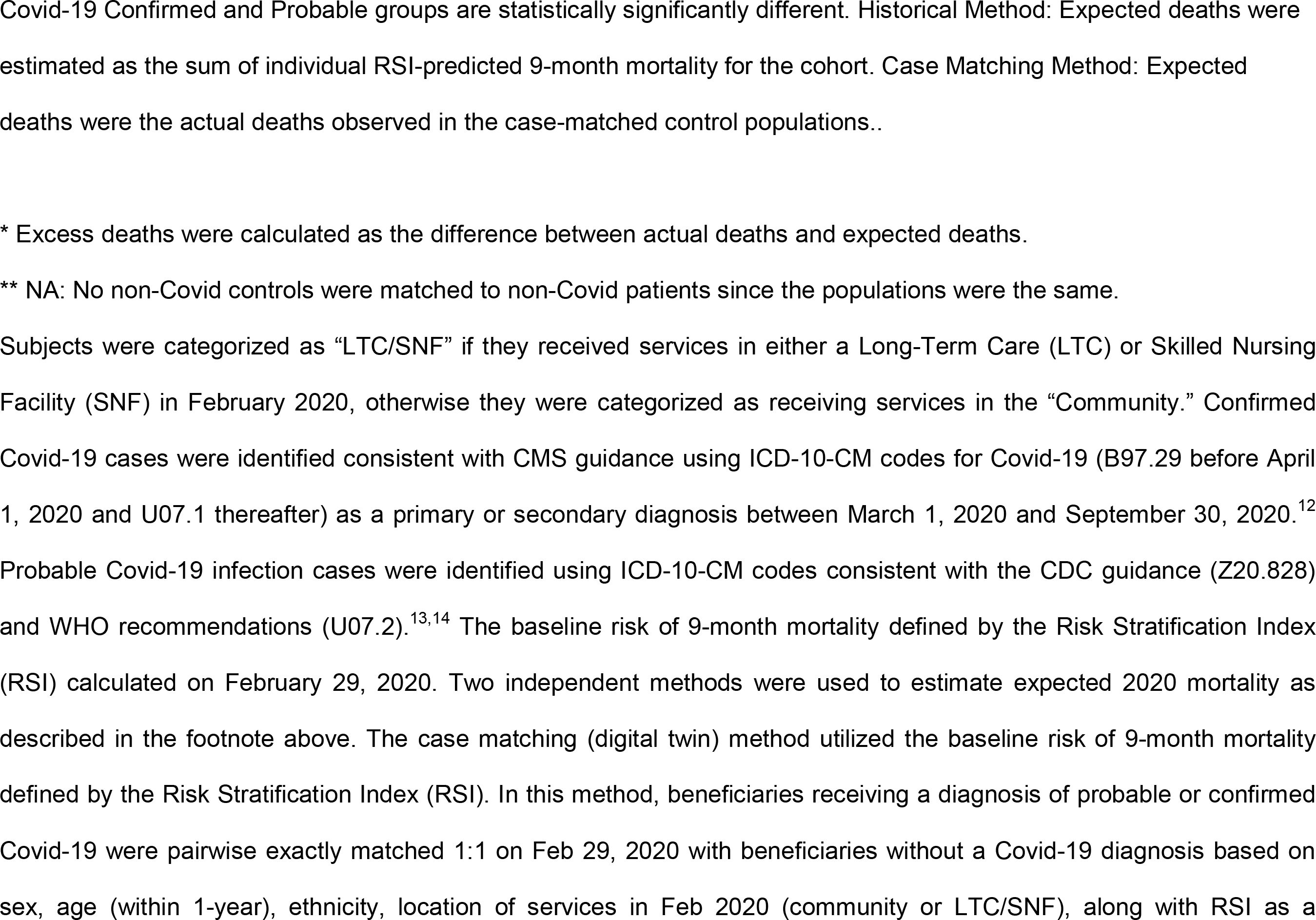
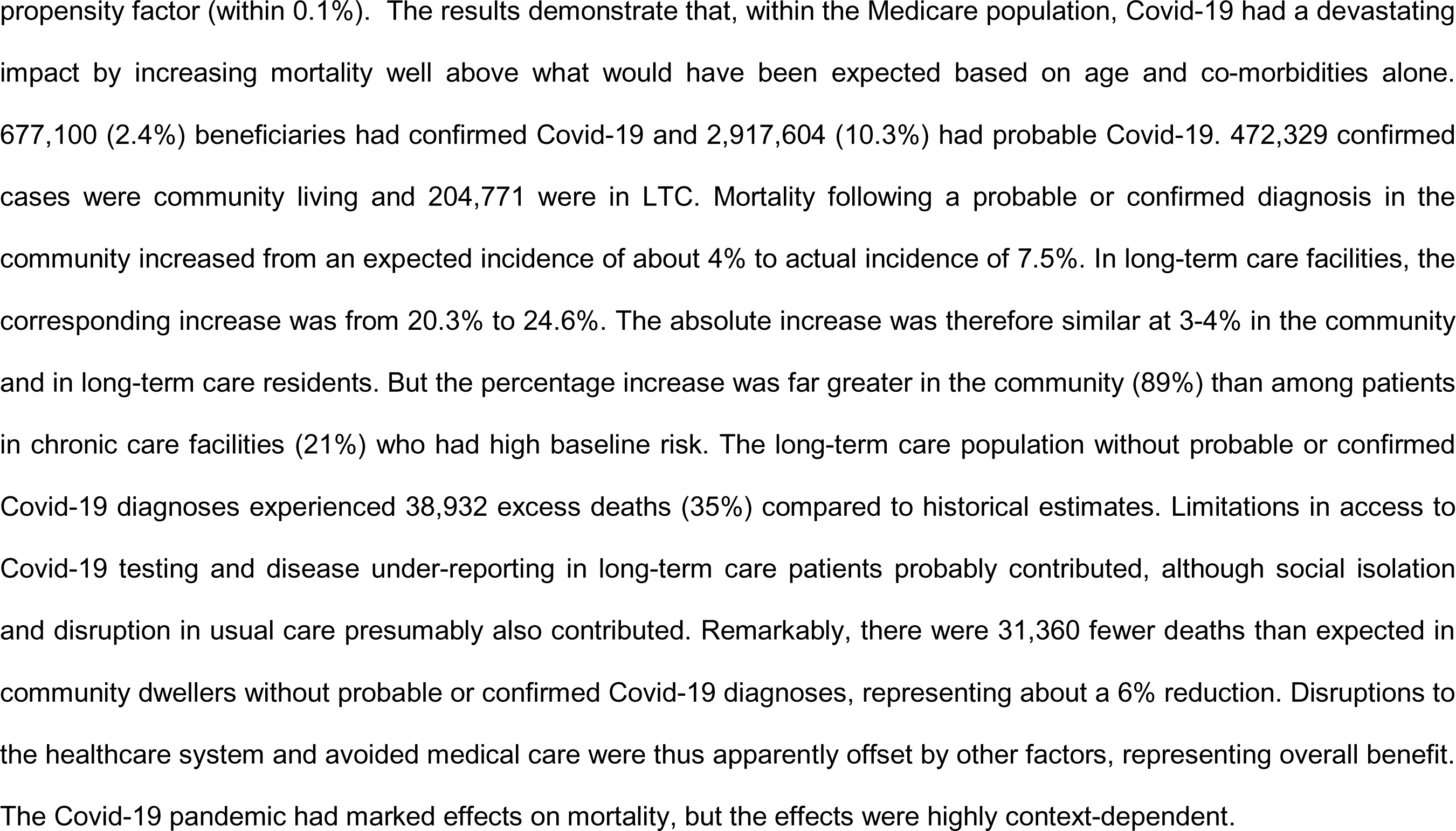
Number of excess deaths by location, diagnosis, and method of calculation.

### Public access

Our model is highly predictive for mortality in Medicare beneficiaries with documented Covid-19 infections. Because baseline RSI scores can help to identify Medicare beneficiaries at highest risk for mortality due to Covid-19, we make the models publicly available in the following formats:

1. Access to RSI risk calculators will be provided free of charge for authorized non-commercial uses via the HDAI API website (https://www.hda-institute.com/api/).
2. Medicare beneficiaries or their health advocates may access their personalized health history and risk assessment by signing into Health Picture (https://my.healthhpicture.com). Health picture is an easy-to-use tool that allows Medicare beneficiaries and their family members a way to access their health histories and understand their Covid-19 risks.
3. Coefficients for a public version of a one-year RSI mortality model are provided at – (Risk Stratification Index | Cleveland Clinic).

## Discussion

Age, sex, care location, and comorbidities were significant predictors of mortality. The strongest individual predictor following a diagnosis of Covid-19 across all age categories, and in both community and long-term care settings was the integrated measure of patient co-morbidities, RSI. Although many individual chronic conditions were also significant risk factors in our unadjusted univariable analysis, the strength of these associations were substantially diminished after adjustment for the primary covariates of age, sex, race, location of services, and RSI. Our analysis is consistent with previous work showing that older, non-white men, and patients receiving services in long term care facilities are at special risk of dying from Covid-19 infections.

Using RSI as a composite measure of baseline mortality risk permitted precise case-control matching, thereby allowing us to estimate excess deaths attributable Covid-19 by two complementary methods in Medicare recipients with probable or confirmed Covid-19 diagnoses. Using the historical comparison, there was an increase from 215,359 expected to 346,062 actual deaths, representing 130,702 excess deaths and a 61% increase. Using matching, mortality increased from 236,119 expected to 337,601 observed deaths, representing 101,482 excess deaths and a 43% increase. Both estimates far exceed the 15-20% excess mortality estimate reported in previous analyses that included younger populations. Our results are therefore consistent with the belief that older people are at much higher risk for developing severe Covid-19 illness — and of dying from it.

Overall, our historical model indicated that mortality following a probable or confirmed diagnosis in the community increased from an expected incidence of about 4% to actual incidence of 7.5%. In LTC/SNF’s, the corresponding increase was from 20.3% to 24.6%. Therefore, the absolute increase in mortality was similar at 3-4% in the community and in long-term care residents. However, baseline risk (RSI) associated with all individuals in a care setting varied greatly, being only about 2.6% in the community versus 20.5% in long-term care facilities. As a percentage, the relative increase in mortality was thus far greater in the community (89%) than among patients in long term care facilities (21.0%).

Somewhat remarkably, overall mortality *decreased* in Medicare participants without probable or confirmed Covid-19 diagnoses. In fact, among community dwellers, there were 31,360 fewer deaths than expected, representing about a 6% reduction. Disruptions to the healthcare system and avoided medical care were thus apparently offset by other factors, representing overall benefit. Obvious health benefits of pandemic isolation include reduced exposure to other airborne illnesses such as influenza, fewer driving accidents and fewer homicides. However, none seems sufficient to explain the reduction. More subtle effects including reduced work or stress-related illness might contribute more, although there is no obvious reason to believe that the pandemic would reduce stress — especially in an over-65-year-old population.

The causes of reduced mortality in community dwelling Medicare participants remains unclear. However, our results suggest that inadequate care for chronic conditions and delayed care of acute events did not produce the feared outcome of higher short-term mortality in the general population without Covid-19. But due to limited follow-up, we caution that disruptions in healthcare delivery may yet result in adverse longer-term outcomes due to delays in the diagnosis and treatment of new and existing chronic conditions. An additional consideration is that prolonged sequela after severe Covid-19 infections (Long Covid syndrome) appear substantial and is an area requiring urgent further study.(18)

There was a distinct disparity between community dwellers and those in long-term care facilities with respect to historical mortality comparisons. In contrast to community Medicare participants, the long-term care population without probable or confirmed Covid-19 diagnoses experienced 38,932 excess deaths (35%) compared to historical estimates. We believe that limitations in access to Covid-19 testing and disease under-reporting in long-term care patients probably were responsible for this finding. It seems likely that many of the excess deaths in this vulnerable population were consequent to undiagnosed Covid-19 infections. But it is also probable that social isolation and disruption in usual care may have contributed as well. The higher-than-expected level of excess deaths observed in this cohort (subjects without a probable or confirmed Covid diagnosis) is reflected in our case matching results, which indicate a modest relative reduction in deaths in subjects with a Covid related diagnosis. This is most likely due to undiagnosed Covid cases included in the control population, but we cannot rule out the possibility that the focus on care for the Covid patients had an unintended adverse impact on the remaining population.

## Limitations

We excluded less than 2.2% of the available population because of missing and inconsistent values. Because data were missing non-systematically, exclusion of these subjects was unlikely to introduce meaningful bias. We relied on administrative diagnostic claims for Covid-19 to assign exposure. Surely these are inexact, especially during our study period early in the pandemic. Furthermore, a new diagnostic code for confirmed Covid-19 (U07.1) was introduced on April 1, 2020, and we assume that there was some uncertainty regarding its proper application. However, Kadri et al recently reported that this Covid-19 specific code showed high sensitivity and specificity compared with the PCR test results.(19) A second limitation is that we did not account for temporal changes in risk of exposure to Covid-19 in either setting, nor for improvements in treatment of infected individuals over time.(20, 21)

We assigned individuals to either community dwelling or long-term care subgroups based on coding in February 2020. Some participants undoubtedly changed their care settings during the analysis period. Skilled nursing facilities, for example, include patients who remain semi-permanently along with others who stay for short periods such during rehabilitation from major orthopedic procedures before resuming community life. But among patients who died, 79% of those who were in a long-term care facility on the anchor date of Feb 29, 2020 had long term care charges within two months of death.

Our analysis was based on 28,389,098 adults enrolled in the US fee-for-service and Medicare/Medicaid program. The results are therefore broadly applicable to Medicare eligible adults. Although our sample included a fair number of dual eligible subjects below age 65, our results should only be cautiously generalized to younger and healthier populations.

## Summary

Mortality following a probable or confirmed Covid-19 diagnosis in the community increased from an expected incidence of about 4% to actual incidence of 7.5%. In long-term care facilities, the corresponding increase was from 20.3% to 24.6%. The absolute increase was therefore similar at 3-4% in the community and in long-term care residents. But the percentage increase was far greater in the community (89%) than among patients in chronic care facilities (21%) who had high baseline risk.

The long-term care population without probable or confirmed Covid-19 diagnoses experienced 38,932 excess deaths (35%) compared to historical estimates. Limitations in access to Covid-19 testing and disease under-reporting in long-term care patients probably contributed, although social isolation and disruption in usual care presumably contributed. Remarkably, there were 31,360 fewer deaths than expected in community dwellers without probable or confirmed Covid-19 diagnoses, representing about a 6% reduction. Disruptions to the healthcare system and avoided medical care were thus apparently offset by other factors, representing overall benefit.

The Covid-19 pandemic had marked effects on mortality, but the effects were highly context-dependent. Among community dwelling Medicare participants with suspected or confirmed Covid-19 diagnoses, mortality nearly doubled, but from a relatively low baseline. Patients in long-term care facilities had a similar absolute increase in mortality, but because their baseline mortality was 20.5%, the relative increase was smaller. In contrast, community dwelling Medicare participants without COVID had about 6% lower-than-expected mortality.

## Author Contributions

Greenwald and Chamoun had full access to all the data in the study and take responsibility for the integrity of the data and the accuracy of the data analysis.

Data Curation: Chamoun, Greenwald

Formal Analysis: Greenwald, Chamoun

Funding Acquisition: Chamoun, Gray

Investigation: Chamoun, Greenwald

Methodology: Manberg, Sessler, Chamoun

Project Administration: Greenwald, Clain

Resources: Chamoun

Software: Chamoun, Greenwald

Supervision: Manberg, Greenwald, Chamoun

Validation: Sessler, Maheshwari, Manberg, Gray, Clain

Visualization: Clain, Greenwald

Writing – Original Draft Preparation: Manberg, Greenwald, Sessler

Writing – Review and Editing: Sessler, Maheshwari, Gray, Clain, Manberg, Chamoun, Greenwald

## Conflict of Interest Disclosures

Greenwald, Chamoun, Manberg, Clain and Gray are employees of and hold equity interest in Health Data Analytics Institute. Dr. Sessler is a paid consultant of and holds equity interest in Health Data Analytics Institute. Dr. Maheshwari received no compensation from Health Data Analytics Institute.

## Funding/Support

The study was funded by Health Data Analytics Institute. The Robert Wood Johnson Foundation provides support for providing access to the Covid-19 RSI_365_ risk prediction models by academic researchers for non-commercial purposes via the HDAI API (https://www.hda-institute.com/api/).

## Role of Funder/Sponsor

Other than the authors and Data Analyst Douceur Tengu, no additional members of Health Data Analytics had a role in the design and conduct of the study; collection, management, analysis, and interpretation of the data; preparation, review, or approval of the manuscript; or decision to submit the manuscript for publication.

We thank John Parks, Zhenyu Hong and Douceur Tengu (Data Analysts, Health Data Analytics Institute) for their assistance in developing models and preparing figures and tables.

## Supporting Information

**S1 Fig. Consort style waterfall flowchart detailing population selection methodology**. Confirmed Covid-19 cases were identified consistent with CMS guidance using ICD-10-CM codes for Covid-19 (B97.29 before April 1, 2020 and U07.1 thereafter) as a primary or secondary diagnosis between March 1, 2020 and September 30, 2020.^12^ Probable Covid-19 infection cases were identified using ICD-10-CM codes consistent with the CDC guidance (Z20.828) and WHO recommendations (U07.2).^13,14^ Subjects were excluded for missing data if values for any baseline characteristic used in the study were missing (i.e., age, sex, ethnicity, location of care, zip code derived measures, dates of coverage, or baseline risk of 9 month mortality assessed with the Risk Stratification Index (RSI).) Additionally, we excluded subjects whose records had inconsistent values among source files containing similar variables such as birth date and sex.

**S2 Fig. Performance characteristics of RSI model. Panel A: ROC curve, Panel B Calibration plot, Panel C Sensitivity and Positive Predicted Value vs probability of mortality**. (A) Area Under the Receiver Operating Curve (AuROC) for the development Learn Set (80% of 2018 Set) was 0.88 (95% Confidence Interval of [0.88-0.88]). AuROC for the prospective Test Set (20% of 2018 Set) was 0.88 (95% Confidence Interval of [0.88-0.88]. Similar performance in the Test Set compared to the Learn Set supports a lack of overfitting in the development of the predictor. (B) The calibration plot displays the mean actual vs predicted 1 year mortality for populations clustered in increments of 1% probability of mortality. Dark green, light green, and red dots are populations of the lowest 95%, 95%-99%, and top 1% risk of mortality. The diagonal line identifies the domain of ideal performance where actual and expected mortality rates are equal for a population. The performance of this index is very close to ideal performance for approximately 99% of the population. Tabulated metrics: The sample size in this test set (N) was 11,923,144 with an incidence of 1yr mortality (Event_Test) of 4.5%. The Slope and Intercept (INT) fit of the data are 0.94 and 0.01, respectively. The area under the Receiver Operating Curve was 0.88. The Mean Average Error (MEA) from cluster coordinates (i.e., (expected, actual) couplets) to the identity line was calculated for the database divided into populations grouped from the riskiest to least risky subjects using cluster sizes ranging from 1 (i.e., each individual as a cluster) to 1000 neighboring subjects (e.g., MAE to MAE_1000). The 95% Confidence Interval (CI) for the fits of these populations to the identify line is tabulated (i.e., AE_CI to AE_CI_1000). Rsq_unit is a goodness of fit measure of individual results to the ideal line.

(C) Positive Predictive Accuracy (blue dots) and Sensitivity (purple dots) versus the fraction of population, sorted by the risk of 1 year mortality. The vertical red line indicates where the number of patients above the risk threshold equals the incidence of mortality in the population. There are several metrics tabulated in the figure: The area under the Receiver Operating Characteristic (ROC) is 0.88. The incidence of mortality (IR) in the population of 11,891,922 was 4.4%. Vertical bars help identify the PPA and sensitivity performance for detectors operating to identify the riskiest 5%, 10%, and 20% patients. The PPV, sensitivity and relative risk (RR) are tabulated for these detector operating points.

**S3 Fig. Comparison of RSI (Panel A) and Chronic Condition based models (Panel B) on 2020 population. ROC curves for all subjects, No Covid-19, Probable Covid-19 and Confirmed Covid-19 populations**. Confirmed Covid-19 cases were identified consistent with CMS guidance using ICD-10-CM codes for Covid-19 (B97.29 before April 1, 2020 and U07.1 thereafter) as a primary or secondary diagnosis between March 1, 2020 and September 30, 2020.^12^ Probable Covid-19 infection cases were identified using ICD-10-CM codes consistent with the CDC guidance (Z20.828) and WHO recommendations (U07.2).^13,14^ (A,B) ROCs display the sensitivity vs. 1 – specificity in detecting patients who died within 9 months after prediction from February 29,2020 (baseline). The areas under each ROC, with their corresponding 95% confidence intervals, are tabulated in the lower right of each figure. Predictions using RSI yielded better performance (A) than those using a model based on age, sex and chronic conditions (B).

**S4 Fig. Forest plot showing the relative risk and 95% CI of significant predictors of confirmed Covid-19 infection**. Confirmed Covid-19 cases were identified consistent with CMS guidance using ICD-10-CM codes for Covid-19 (B97.29 before April 1, 2020 and U07.1 thereafter) as a primary or secondary diagnosis between March 1, 2020 and September 30, 2020.^12^ Subjects were categorized as “LTC/SNF” if they received services in either a Long Term Care (LTC) or Skilled Nursing Facility (SNF) in February 2020, otherwise they were categorized as receiving services in the “Community.” Predictors were assessed at baseline (February 29, 2020) and include quintiles of Risk Stratification Index (RSI), presence of chronic conditions, location of services (LTC/SNF vs Community), and demographic variables (i.e., age, sex, race, and quintiles of median household income imputed by zip code according to 2015 Census data.) Variables not remaining in the adjusted model are indicated by the presence of empty parenthesis under the adjusted odds ratio. Location of services, age, status of end-stage renal disease (ESRD) and RSI were the strongest (unadjusted) predictors of infection. Location of services and ESRD remained strong predictors following adjustment; however, risks associated with having chronic conditions were typically reduced when adjusted by the presence of other factors.

**S5 Fig. Observed mortality rates by age and RSI quintiles**. Rates of mortality within 9 months following baseline (February 29, 2020) in Medicare subpopulations categorized by age, location of services, infection status, and quintiles of the baseline risk of mortality assessed using the Risk Stratification Index (RSI). Confirmed Covid-19 cases were identified consistent with CMS guidance using ICD-10-CM codes for Covid-19 (B97.29 before April 1, 2020 and U07.1 thereafter) as a primary or secondary diagnosis between March 1, 2020 and September 30, 2020.^12^ Probable Covid-19 infection cases were identified using ICD-10-CM codes consistent with the CDC guidance (Z20.828) and WHO recommendations (U07.2).^13,14^ Subjects were categorized as “LTC/SNF” if they received services in either a Long-Term Care (LTC) or Skilled Nursing Facility (SNF) in February 2020, otherwise they were categorized as receiving services in the “Community.” As expected, subjects in quintiles with higher baseline risk of mortality had higher rates of observed mortality. For subjects without a Covid diagnosis, mortality rates were lower in the community setting compared to those in the LTC/SNF; however, for subjects with confirmed or probable Covid infection, mortality rates were typically higher in the community setting than in the LTC/SNF.

**S1 File. RSI Development Method for 9-month predictions**.

## Notes

### Clinical Trial

This manuscript reports outcomes of a retrospective study on previously collected data by the Centers of Medicare and Medicaid.

### Author Declarations

This project was determined to be exempt from informed consent requirements by the New England Institutional Review Board (https://www.wcgirb.com/). This research qualifies under the approved exempt category: "Research involving the collection or study of existing data, documents, records, pathological specimens, or diagnostic specimens, if the investigator records the information in such a manner that subjects cannot be identified, directly or through identifiers linked to the subjects."

## References

1. CDC. CDC COVID Data Tracker: Centers for Disease Control and Prevention; 2021 [Available from: https://covid.cdc.gov/covid-data-tracker/#cases_casesper100klast7days.

2. Chen YH, Glymour MM, Catalano R, Fernandez A, Nguyen T, Kushel M, et al. Excess Mortality in California During the Coronavirus Disease 2019 Pandemic, March to August 2020. JAMA Intern Med. 2020.

3. Faust JS, Krumholz HM, D. C, Mayes KD, Lin Z, Gilman C, et al. All-Cause Excess Mortality and COVID-19–Related Mortality Among US Adults Aged 25-44 Years, March-July 2020. JAMA. 2020.

4. Lauren M. Rossen AMB, Farida B. Ahmad, Paul Sutton, Robert N. Anderson. Excess Deaths Associated with COVID-19, by Age and Race and Ethnicity — United States, January 26–October 3, 2020 2021 [Available from: https://www.cdc.gov/mmwr/volumes/69/wr/mm6942e2.htm.

5. CDC. Medical Conditions and Risk of for Severe COVID-19 Illness: Centers for Disease Control and Prevention; 2021 [Available from: https://www.cdc.gov/coronavirus/2019-ncov/need-extra-precautions/people-with-medical-conditions.html.

6. Wang Q, Davis PB, Gurney ME, Xu R. COVID-19 and dementia: Analyses of risk, disparity, and outcomes from electronic health records in the US. Alzheimers Dement. 2021.

7. Harrison SL, Fazio-Eynullayeva E, Lane DA, Underhill P, Lip GYH. Comorbidities associated with mortality in 31,461 adults with COVID-19 in the United States: A federated electronic medical record analysis. PLoS Med. 2020;17(9):e1003321.

8. Wang Q, Berger NA, Xu R. Analyses of Risk, Racial Disparity, and Outcomes Among US Patients With Cancer and COVID-19 Infection. JAMA Oncol. 2021;7(2):220–7.

9. Wynants L, Van Calster B, Collins GS, Riley RD, Heinze G, Schuit E, et al. Prediction models for diagnosis and prognosis of covid-19 infection: systematic review and critical appraisal. BMJ. 2020;369:m1328.

10. Experton B, Tetteh HA, Lurie N, Walker P, Carroll CJ, Elena A, et al. A Predictive Model for Severe Covid-19 in the Medicare Population: A Tool for Prioritizing Scarce Vaccine Supply. medRxiv. 2020:2020.10.28.20219816.

11. Dun C, Walsh CM, Bae S, Adalja A, Toner E, Lash TA, et al. A Machine Learning Study of 534,023 Medicare Beneficiaries with COVID-19: Implications for Personalized Risk Prediction. medRxiv. 2020:2020.10.27.20220970.

12. von Elm E, Altman DG, Egger M, Pocock SJ, Gotzsche PC, Vandenbroucke JP, et al. The Strengthening the Reporting of Observational Studies in Epidemiology (STROBE) statement: guidelines for reporting observational studies. Lancet. 2007;370(9596):1453–7.

13. CMS. Preliminary Medicare COVID-19 Data Snapshot CMS: Centers for Medicare & Medicaid Services; 2021 [Available from: https://www.cms.gov/research-statistics-data-systems/preliminary-medicare-covid-19-data-snapshot.

14. CDC. ICD-10-CM Official Coding and Reporting Guidelines April 1, 2020 through September 30, 2020. Centers for Disease Control and Prevention; 2020.

15. WHO. COVID-19 coding in ICD-10: World Health Organization; 2020 [cited World Health Organization. Available from: https://www.who.int/classifications/icd/COVID-19-coding-icd10.pdf.

16. Chamoun GF, Li L, Chamoun NG, Saini V, Sessler DI. Validation and Calibration of the Risk Stratification Index. Anesthesiology. 2017;126(4):623–30.

17. CMS. Condition Categories - Chronic Conditions Data Warehouse: Centers for Medicare & Medicaid Services; 2021 [Available from: https://www2.ccwdata.org/web/guest/condition-categories.

18. Lopez-Leon S, Wegman-Ostrosky T, Perelman C, Sepulveda R, Rebolledo PA, Cuapio A, et al. More than 50 Long-term effects of Covid-19: a systematic review and meta-analysis. medRxiv preprint. 2021.

19. Kadri SS, Gundrum J, Warner S, Cao Z, Babiker A, Klompas M, et al. Uptake and Accuracy of the Diagnosis Code for COVID-19 Among US Hospitalizations. JAMA. 2020;324(24):2553–4.

20. Nguyen NT, Chinn J, Nahmias J, Yuen S, Kirby KA, Hohmann S, et al. Outcomes and Mortality Among Adults Hospitalized With COVID-19 at US Medical Centers. JAMA Netw Open. 2021;4(3):e210417.

21. Tenforde MW, Fisher KA, Patel MM. Identifying COVID-19 Risk Through Observational Studies to Inform Control Measures. JAMA. 2021.

