## Supplementary figures and images for "Covid-19 and Excess Mortality in Medicare Beneficiaries"

### S1 Fig

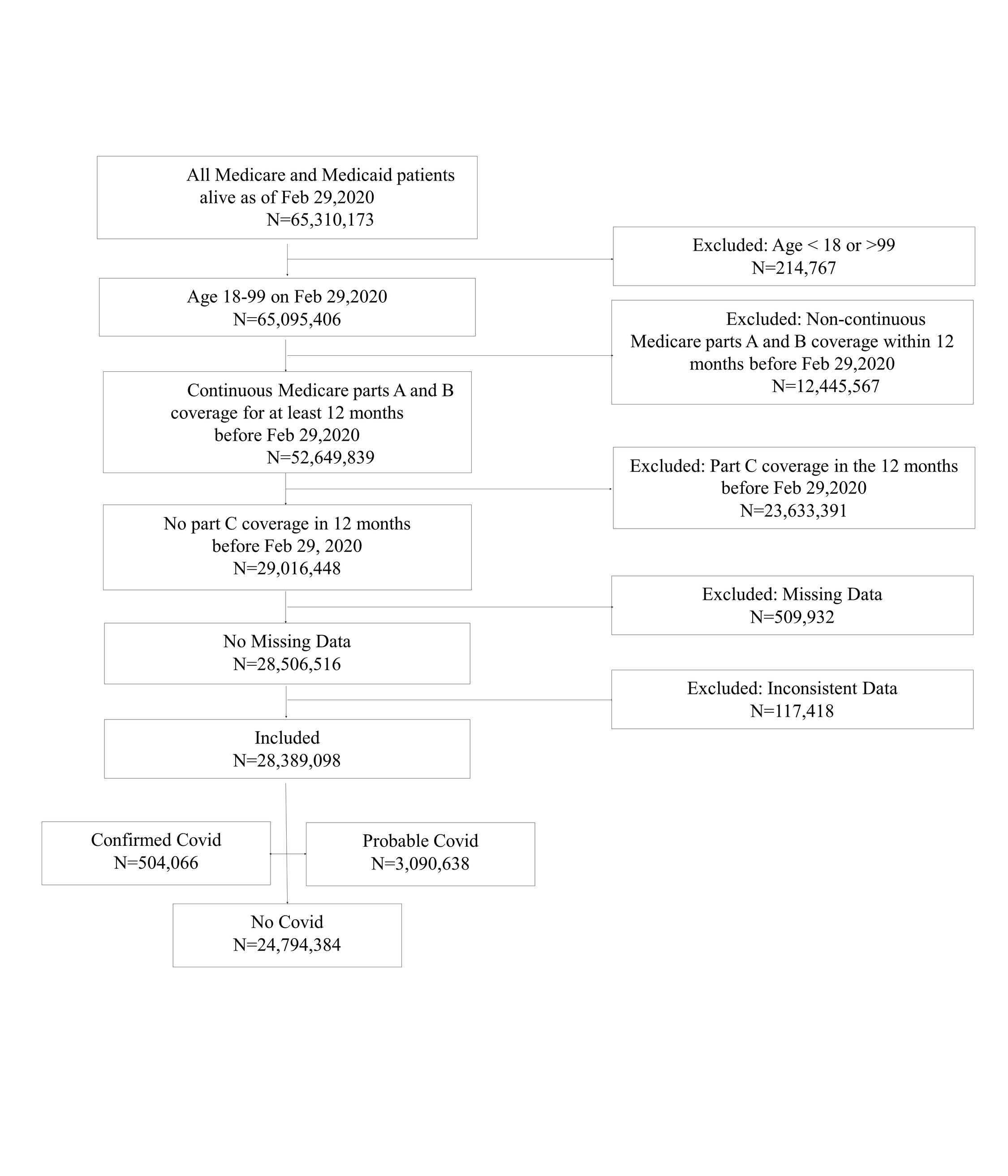

### S2 Fig

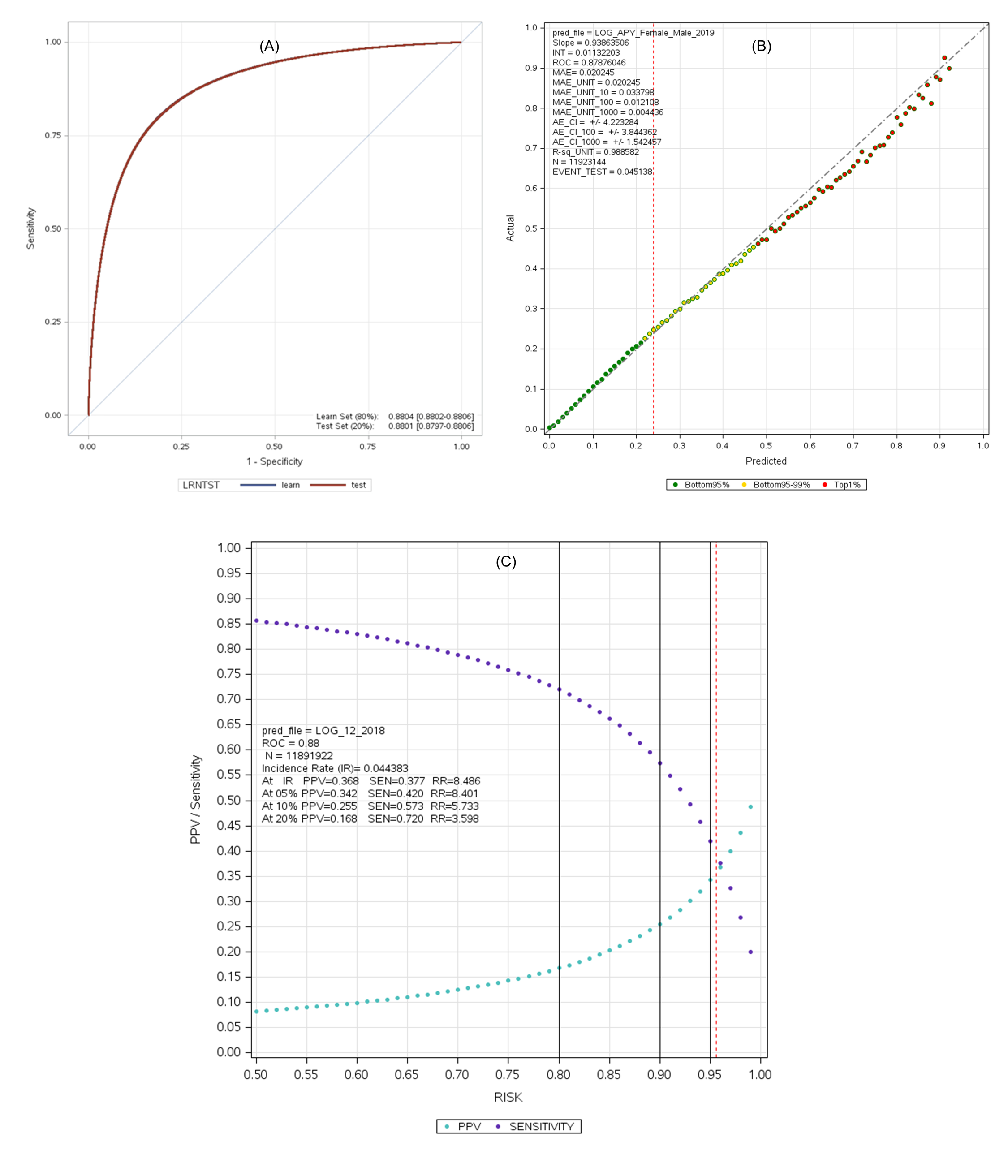

### S3 Fig

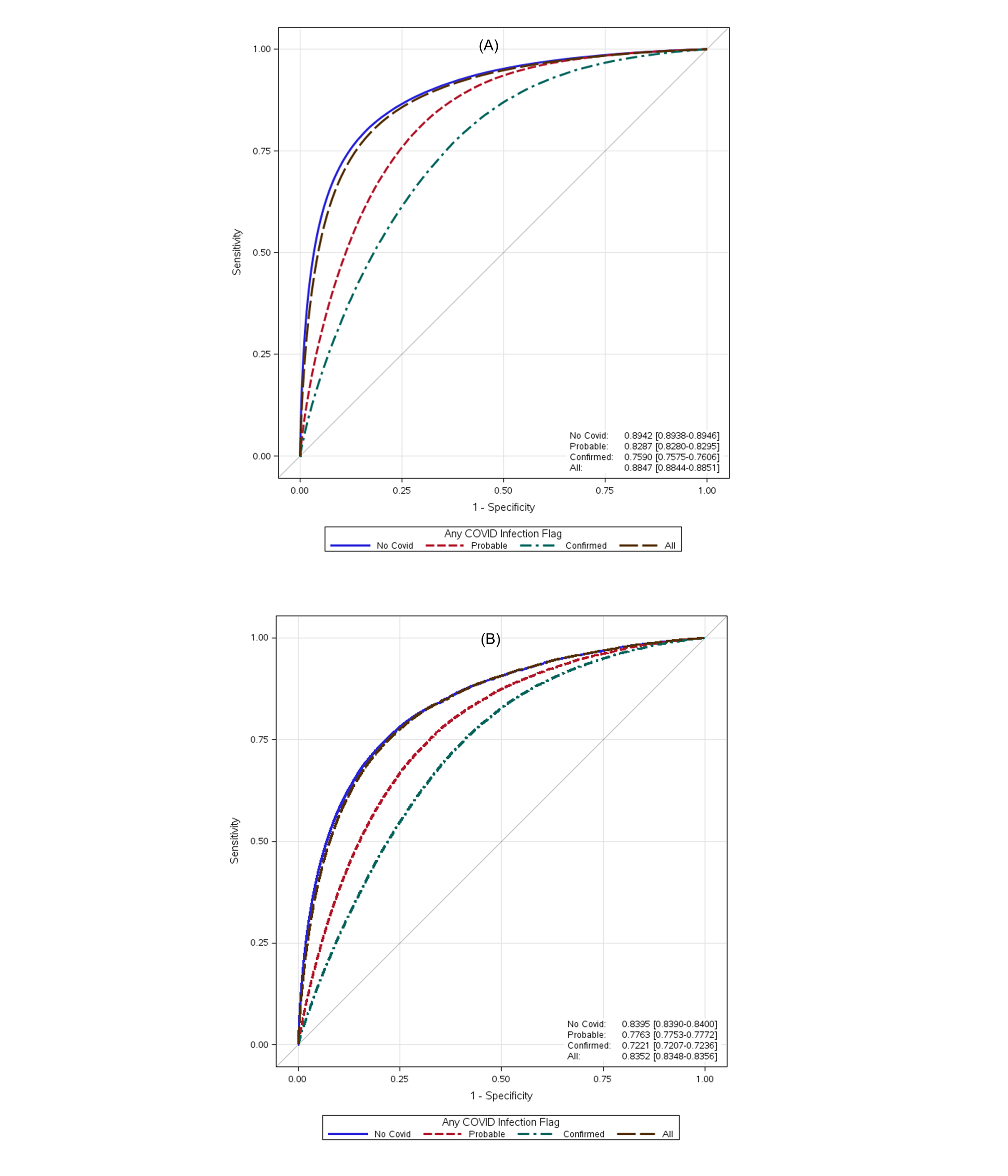

### S4 Fig

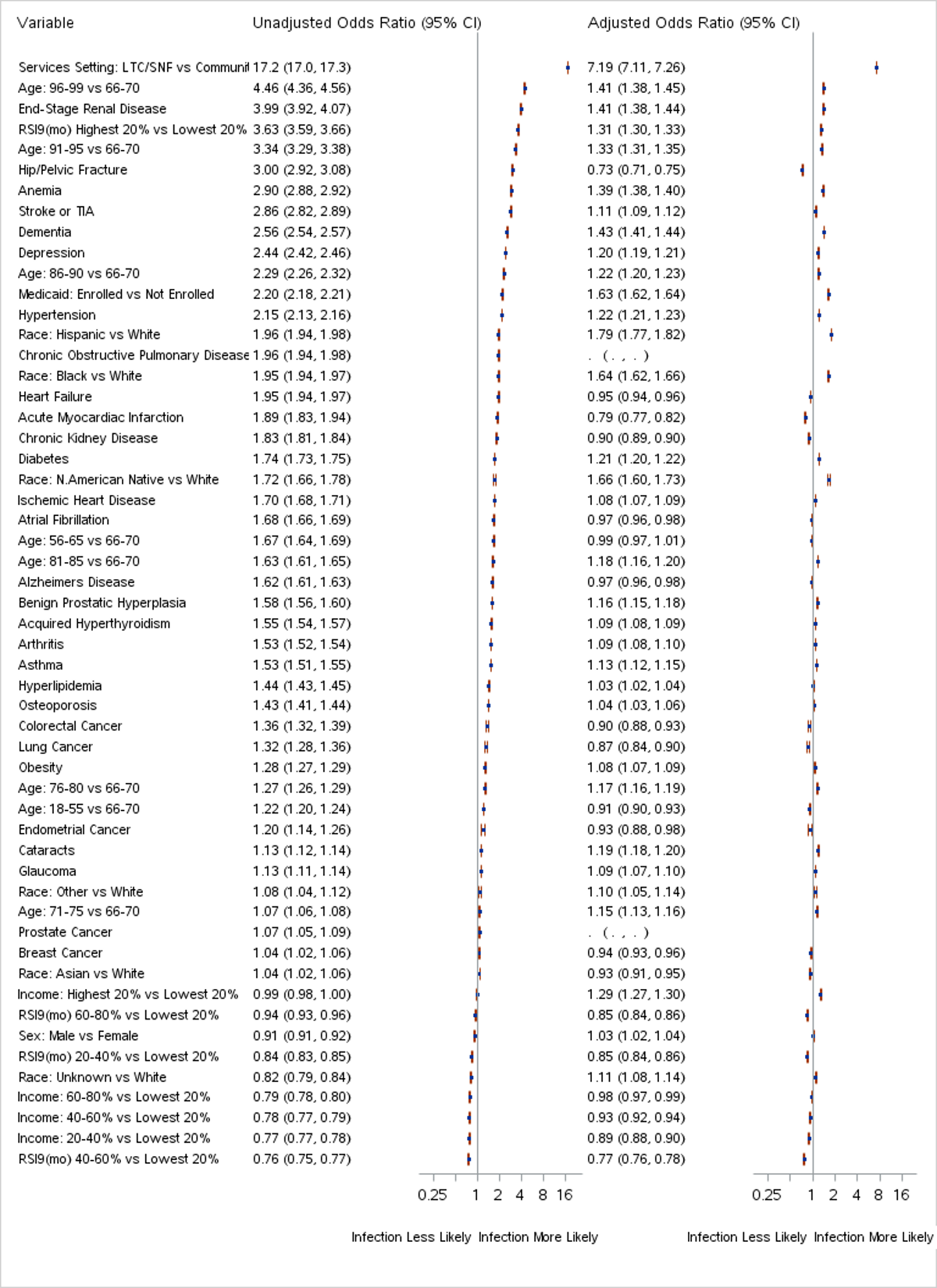

### S5 Fig

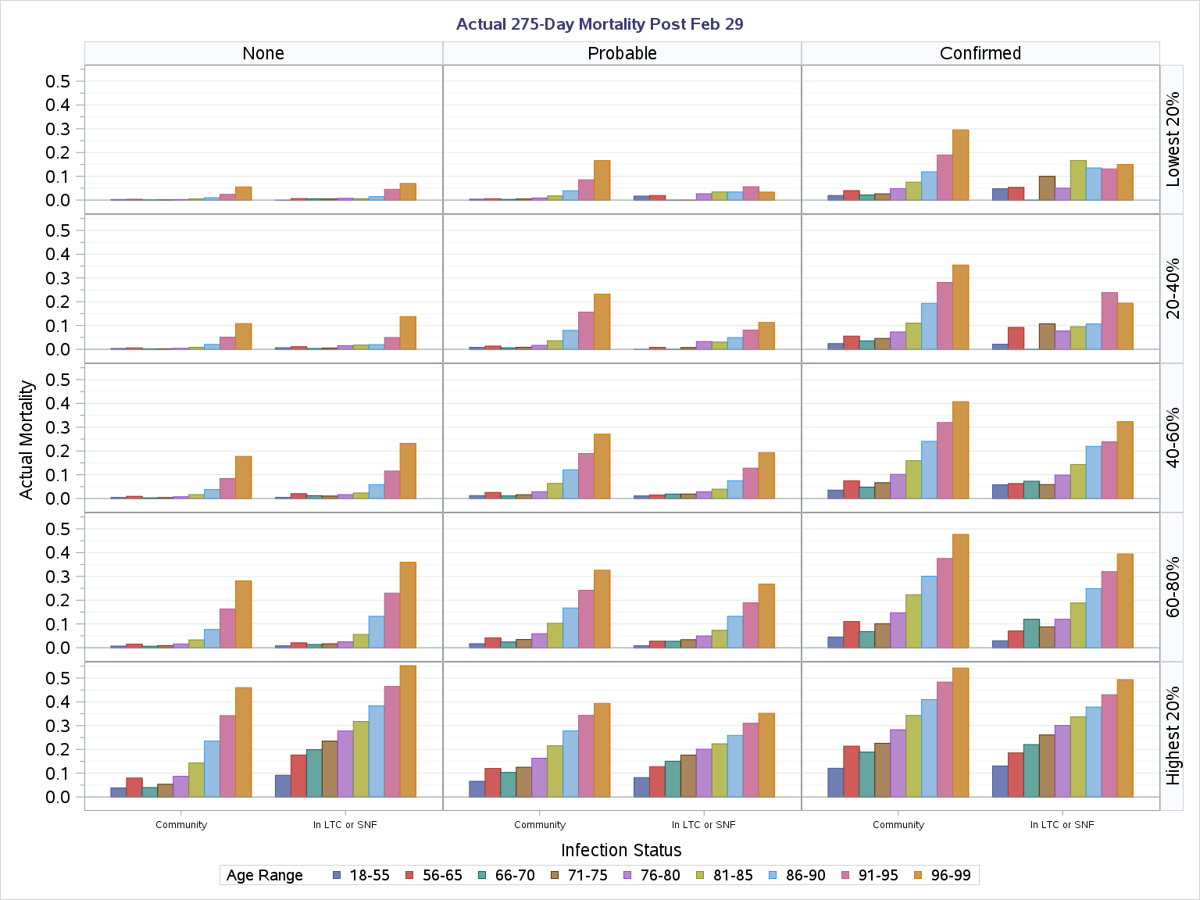
