## Supplementary material for "Covid-19 and Excess Mortality in Medicare Beneficiaries": S1 File

**S1 File. RSI Development Method for 9-month predictions.**

Models to predict 1 year mortality were developed first for each calendar year (through 2018) in the following manner:

First, for purposes of model development, each patient was randomly assigned to one day in the year which serves as his/her reference date (i.e., “anchor date”) from which mortality is predicted. The patient is assigned to the same day in each year for all years the patient is available. Randomization to dates within a year provides 365 independent - but similar- cohorts (i.e., the distribution of sex, age, chronic conditions, etc. are similar among the groups.) This construct has 2 benefits: 1) it permits comparisons of outcomes among similar populations at different time points throughout the year (enabling analyses of seasonal influences), and 2) it permits year-over-year longitudinal analyzes within each population. During the development process, no patient is assigned an anchor date of Feb 29$^{th}$; however, once a model is developed, the coefficients can be applied to any date of the year, including Feb 29$^{th}$.

Second, the medical status of each patient on the anchor date was compiled from claim records and represented by a series of binary variables cataloging the presence (1) or absence (0) of a set of predefined ICD medical and surgical codes observed within the year prior to the patient’s anchor date. The general method used to identify the set of predefined ICD codes was previously described[1].

Third, the mortality status of each patient one year forward from their anchor date was derived from the recorded date of death or its absence. The combined set of variables representing the medical status, sex, age, Medicaid enrollment (dual eligibility) status and mortality outcome for all patients for a given year of anchor dates is called an “annual dataset”.

Finally, logistic regression (stepwise selection) was used to create a model to predict 1 year mortality using two sequential years of annual datasets as the development database (i.e., the current and past year’s datasets.) For example, the “2018” model was derived from data associated with anchor dates in 2017 and 2018 predicting mortality outcomes observed in 2018 and 2019. For each model derivation, the development database was partitioned into a learning set (80% of subjects) for model generation and a prospective evaluation set (20% of subjects). Example performance results of this process for the 2018 model are shown above. The receiver operating curve (ROC) characteristics shown in **S2 Fig Panel A** demonstrates the lack of overfitting using this methodology because the ROC curve of the prospective set is nearly identical to the ROC curve of the development set. **S2 Fig Panel B** illustrates the calibration characteristic for the prospective set, analyzed at one percent resolution of risk of mortality. These results demonstrate a tight match between the actual and expected mortality of groups of patients along the continuum of risk for 99% of the population (i.e., dark and light green populations). **S2 Fig Panel C** shows the relationship of sensitivity and positive predictive accuracy with the predicted risk of mortality.

Predictions of 9-month mortality were derived using a three-step calibration process from predictions of 1yr mortality generated from 3 annual models. The following example describes predictions of 9mo mortality for all 2020 subjects from Feb 29,2000.

First, the 2018 annual 1 year mortality model (described above) was applied to the 2020 cohort in which the patient medical history and mortality outcome were referenced to an anchor date of Feb 29, 2020 for each patient in the dataset. This provided an initial estimate of 1 year mortality for the 2020 cohort (i.e., MORX0365_P).

To create a mapping construct between the predicted 1-year mortality and the 9mo mortality, bins of 0.1% resolution of risk (i.e., MORX0365_P_P001) were derived from MORX0365_P by truncating the prediction in 0.001 increments. Subsequently, 9mo-mortality status for each patient was determined for the 3 annual datasets prior to 2020. Nine-month mortality outcomes were then pooled for each 0.1% bin for each day of a given year.

To improve the estimation of actual 9-mo mortality on a given date within a year, estimated actual mortality for a given date was derived from the 60-day centered moving average of raw daily mortality counts. Specifically, the daily estimate of actual mortality per bin was derived from the ratio of the sum of mortality count within the 60-day window [numerator] divided by the sum of the number of observations within the 60-day window [denominator]. This averaging process provides a robust estimate of daily mortality per bin of risk, improving calibration performance particularly in low-risk populations. Subsequently, the initial estimate of actual 9mo mortality for a date in the future (e.g., 2020) was derived from combining estimates of actual mortality on the same day of the year from the annual datasets of the prior 3 years. Specifically, for each of the 1000 risk bins per day in 2020, the "predicted 9mo mortality" was calculated as the ratio of the sum of the 3 numerators divided by the sum of the 3 denominators from the prior 3 years.

Finally, to smooth transitions of predicted 9-mo mortality risk among adjacent risk bins, a third-order spline was fit to the 1000 daily pairs of initial estimates of 9-mo and predicted 1yr mortality. The spline-predicted 9mo mortality estimates then provided the final calibration map per day between bins of 1yr risk of mortality (created from the 2018 model applied to 2020 data) and the predicted 9mo risk of mortality (estimated from the prior 3yrs of experience.)

1. Chamoun GF, Li L, Chamoun NG, Saini V, Sessler DI. Validation and Calibration of the Risk Stratification Index. *Anesthesiology.* 2017;126(4):623-630.
